# Proteomic Networks and Related Genetic Variants Associated with Smoking and Chronic Obstructive Pulmonary Disease

**DOI:** 10.1101/2024.02.26.24303069

**Authors:** Iain R Konigsberg, Thao Vu, Weixuan Liu, Elizabeth M Litkowski, Katherine A Pratte, Luciana B Vargas, Niles Gilmore, Mohamed Abdel-Hafiz, Ani W Manichaikul, Michael H Cho, Craig P Hersh, Dawn L DeMeo, Farnoush Banaei-Kashani, Russell P Bowler, Leslie A Lange, Katerina J Kechris

## Abstract

**Background:** Studies have identified individual blood biomarkers associated with chronic obstructive pulmonary disease (COPD) and related phenotypes. However, complex diseases such as COPD typically involve changes in multiple molecules with interconnections that may not be captured when considering single molecular features.

**Methods:** Leveraging proteomic data from 3,173 COPDGene Non-Hispanic White (NHW) and African American (AA) participants, we applied sparse multiple canonical correlation network analysis (SmCCNet) to 4,776 proteins assayed on the SomaScan v4.0 platform to derive sparse networks of proteins associated with current vs. former smoking status, airflow obstruction, and emphysema quantitated from high-resolution computed tomography scans. We then used NetSHy, a dimension reduction technique leveraging network topology, to produce summary scores of each proteomic network, referred to as NetSHy scores. We next performed genome-wide association study (GWAS) to identify variants associated with the NetSHy scores, or network quantitative trait loci (nQTLs). Finally, we evaluated the replicability of the networks in an independent cohort, SPIROMICS.

**Results:** We identified networks of 13 to 104 proteins for each phenotype and exposure in NHW and AA, and the derived NetSHy scores significantly associated with the variable of interests. Networks included known (sRAGE, ALPP, MIP1) and novel molecules (CA10, CPB1, HIS3, PXDN) and interactions involved in COPD pathogenesis. We observed 7 nQTL loci associated with NetSHy scores, 4 of which remained after conditional analysis. Networks for smoking status and emphysema, but not airflow obstruction, demonstrated a high degree of replicability across race groups and cohorts.

**Conclusions:** In this work, we apply state-of-the-art molecular network generation and summarization approaches to proteomic data from COPDGene participants to uncover protein networks associated with COPD phenotypes. We further identify genetic associations with networks. This work discovers protein networks containing known and novel proteins and protein interactions associated with clinically relevant COPD phenotypes across race groups and cohorts.

## Introduction

In the US, chronic obstructive pulmonary disease (COPD) is a major public health concern as the fourth leading cause of death [1], affecting more than 16 million adults [2]. COPD is characterized by lung inflammation and the diagnosis of chronic airflow obstruction is made using spirometry [3]. Tobacco smoking is the primary exposure risk factor for the development of COPD in the US. Staudt et al. [4] showed that tobacco smoke diminished the capacity to regenerate airway epithelium in COPD. It is not unexpected, therefore, that 42.3% of current and former smokers with normal spirometry [5] have respiratory symptoms and evidence of emphysema or airway thickening on chest computed tomography (CT) scans.

Forced expiratory volume in one second (FEV_1_) and percent emphysema (%LAA950) are clinically observable characteristics related to symptoms, exacerbations, and response to treatment [6]. Being a non-invasive, inexpensive, highly accessible, and easily reproducible method, spirometry is the current gold standard for diagnosing and monitoring COPD progression [7]. Emphysema is another phenotype of COPD, which describes obliteration of the acinar units of the lung [8]. Emphysema can be quantified by lung density measured from CT images in which dense lung tissue is replaced by less dense air [9].

Recent advances in high throughput technologies allow investigators to collect data from multiple biological layers including the genome, transcriptome, and metabolome [10–12]. In particular, the proteome, where peptide and protein abundance are quantified, has posed a great advantage in studying complex diseases such as COPD since proteins play direct functional roles in biological systems and may provide more relevant information related to disease mechanisms than transcriptional profiling [13]. Previous studies have focused on individual proteins associated with COPD [14]. Lee et al. [15] identified eight up-regulated proteins in the COPD group in comparison with the nonsmoker group. Similarly, Ohlmeier et al. [16] observed increased levels of surfactant protein A (SP-A) in COPD participants but not in the normal or fibrotic lung by investigating changes in the proteome from human lung tissue.

While these studies identified individual protein biomarkers with prognostic potential, they were limited by small sample sizes in hard-to-obtain lung tissue and lack the additional predictive power gained by simultaneously considering a collection of related biomarkers and their interactions [17]. Consequently, network-based analyses have emerged as a powerful framework to characterize changes in multiple molecular entities and their interconnections that may not be captured by single molecular features [18]. Obeidat et al. [19] constructed networks of co-expressed genes from peripheral blood of COPD patients using weighted correlation network analysis (WGCNA) [20] and identified networks associated with forced expiratory volume in one second (FEV_1_) and enriched in interleukin (IL)-10 and IL-8 signaling pathways. In another study, Mammen et al. [21] performed network analysis on proteomic data collected from bronchoalveolar lavage of the epithelial lining fluid (BALF) samples and identified 233 differentially expressed proteins in moderate COPD compared to controls. Topological analysis of these proteins suggested the importance of intercellular adhesion molecule 1 (ICAM1), galectin-3, fibronectin, and vimentin in mediating inflammation and fibrogenesis.

Most large-scale omics studies for COPD have been conducted in primarily European ancestry populations while only a limited number of relatively smaller-sized studies have focused on other populations [15, 22–26]. Polygenic risk scores (PRS) provide complementary information for predicting COPD and related phenotypes [27], however, they present a large amount of uncertainty which limits the transferability across ancestry groups [28, 29]. Motivated by the lack of COPD omics studies in non-European ancestries, we conducted proteomic analyses on a large cohort that includes >1,500 self-described African American (AA) subjects to gain more insights into potential proteomic signatures associated with the disease. We leverage proteomic data and network-based approaches to identify protein networks associated with COPD phenotypes separately in AA and Non-Hispanic White (NHW) participants.

In this work, we used sparse multiple canonical correlation network analysis (SmCCNet) [30] to construct proteomic networks associated with two COPD phenotypes (FEV_1_ and emphysema quantified as percentage of low-attenuation areas defined by voxels with Hounsfield Units < 950 (%LAA950)) and a relevant exposure (current smoking status) across two race groups (AA and NHW).

The resulting networks were compared to identify common, phenotype- and race-group-specific proteins and their corresponding interactions to gain insights into the underlying mechanisms of COPD. As proteins can have strong genetic associations [31–33] that may reflect upstream regulatory process, we also performed a genome-wide quantitative trait loci (QTL) analysis to identify loci associated with each network, in addition to QTL analyses of individual proteins in the networks. Through colocalization and conditional analyses, we further investigated whether the genetic associations observed were due to individual effects of the proteins in the network versus a cumulative effect of the network. Finally, we demonstrated that networks for smoking and %LAA950 built in one race group generally transfer to the other and that networks also validated in an external cohort, the SubPopulations and InteRmediate Outcome Measures in COPD Study (SPIROMICS).

## Materials and Methods

### COPD Cohorts

*COPDGene* [34] (Clinical Trial Registration NCT02445183) is a large, multi-center observational study that enrolled 10,198 current and former smokers with at least a 10 pack-year history of smoking, as well as additional never smoker controls (< 100 lifetime cigarettes) with and without COPD, 45-80 years old, with 2/3 non-Hispanic white and 1/3 African Americans. Genotyping data were from the enrollment visit. Proteomics was generated at the five-year follow-up (2013 and 2017, Visit 2) [34] [35] (**Supplement Figure 1**). All study participants provided informed written consent.

*SPIROMICS* (Clinical Trial Registration NCT01969344) is a multi-center observational study that enrolled 2,973 current and former smokers with at least 20 pack-years of smoking between November 2011 to January 2015. Subject were between 40-80 years of age at the time of enrollment and were categorized into never-smokers (<1 pack-year, Stratum 1) or history of smoking (>20 pack years) and divided by spirometry into strata; Stratum 2: FEV1/FVC > 0.7 and FVC > LLN; Stratum 3 : FEV1/FVC <0.07 and FEV1>50% predicted; Stratum 4: FeV1/FVC <0.07 and FEV1‘<50% predicted). The cohort is multiracial with 73% non-Hispanic white, 18% African American, and 9% other races. Fasting blood was collected at visit 1 in vacutainer EDTA plasma tube, immediately spun, aliquoted, frozen, and stored at –80°C [36]. For replication we used the smokers (strata 2-4) non-Hispanic NHW and African American race groups at Visit 1 who had SomaScan v 4.1 profiles (n= 1792). All study participants provided informed written consent (**Supplement Table 2, Supplement Figure 2**).

### COPDGene Cohort Demographics

Proteomic analyses included 3,173 COPDGene participants. Demographics and relevant clinical characteristics of participants, stratified by self-identified race, are shown in **Table 1**. All participants are current or former smokers. We applied a matching approach in an attempt to better match the NHW and AA groups in terms of age, smoking status, sex, and GOLD stage (see **Supplement Table 1** and **Table 1** for details). Further details on matching are in **Supplementary Methods.**

**Table 1.** Characteristics of COPDGene matched study populations and SPIROMICS by race.

|  | COPDGene |  | SPIROMICS |  |
| --- | --- | --- | --- | --- |
| Characteristics | NHW<br>N=1660 | AA<br>N=1513 | NHW<br>N=1459 | AA<br>N=333 |
| **Demographics** |  |  |  |  |
| Age (yr) mean (SD) | 62.9 (8.0)* | 60.2 (7.1)* | 65.42 (8.18)<sup>£</sup> | 58.46 (8.71)<sup>£</sup> |
| Males n(%) | 790 (47.6%) | 745 (49.2%) | 650 (44.6)<sup>£</sup> | 175 (52.6)<sup>£</sup> |
| Females n(%) | 870 (52.4%) | 768 (50.8%) | 809 (55.4)<sup>£</sup> | 158 (47.4)<sup>£</sup> |
| **Smoking Exposure** |  |  |  |  |
| Smoking Status n(%): Former | 647 (39%) | 531 (35.1%) | 965 (67.2)<sup>£</sup> | 116 (35.2)<sup>£</sup> |
| Current | 1013 (61.0%) | 982 (64.9) | 470 (32.8)<sup>£</sup> | 214 (64.8)<sup>£</sup> |
| Pack-years median(IQR) | 42.3 (25.5)* | 35.7 (24.3)* | 45.00 (27.0)<sup>£</sup> | 37.50 (20.0)<sup>£</sup> |
| **Clinical** |  |  |  |  |
| BMI kg/m<sup>2</sup> (mean(SD)) | 28.4 (6.3)* | 29.4 (7.1)* | 27.77 (5.09) | 27.67 (6.06) |
| COPD GOLD Stages n(%): PRISm | 209 (12.6%)* | 256 (16.9%)* | 33 (2.3)<sup>£</sup> | 13 (3.9)<sup>£</sup> |
| GOLD 0 Smoker Controls | 678 (40.8%)* | 727 (48.1%)* | 389 (26.7)<sup>£</sup> | 145 (43.7)<sup>£</sup> |
| GOLD 1 | 158 (9.5%)* | 105 (6.9%)* | 237 (16.3)<sup>£</sup> | 32 (9.6)<sup>£</sup> |
| GOLD 2 | 370 (22.3%)* | 247 (16.3%)* | 465 (31.9)<sup>£</sup> | 70 (21.1)<sup>£</sup> |
| GOLD 3 | 185 (11.1%)* | 129 (8.5%)* | 235 (16.1)<sup>£</sup> | 47 (14.2)<sup>£</sup> |
| GOLD 4 | 60 (3.6%)* | 49 (3.2%)* | 97 (6.7)<sup>£</sup> | 25 (7.5)<sup>£</sup> |
| **Pulmonary Function** (mean(SD)) |  |  |  |  |
| FEV<sub>1</sub> (liter) | 2.3 (0.9)* | 2.1 (0.8)* | 2.09 (0.90) | 1.99 (0.87) |
| FEV<sub>1</sub> Percent Predicted | 76.8 (23.4)* | 79.9 (23.7)* | 71.88 (25.87)<sup>£</sup> | 76.32 (28.06)<sup>£</sup> |
| FEV<sub>1</sub>/FVC | 0.67 (0.15)* | 0.71 (0.14)* | 0.59 (0.16)<sup>£</sup> | 0.64 (0.17)<sup>£</sup> |
| FVC (liter) | 3.3 (1.0)* | 2.9 (0.9)* | 3.51 (1.02)<sup>£</sup> | 3.05 (0.93)<sup>£</sup> |
| **Emphysema** |  |  |  |  |
| Emphysema (% LAA < -950 HU) median(IQR) | 1.5 (4.4)* | 0.9 (3.4)* | 3.84 (10.23)<sup>£</sup> | 1.78 (10.33)<sup>£</sup> |
| PD15<sub>adj</sub> (g/L) | 87.0 (24.0)* | 95.1 (28.0)* | 78.91 (25.42)<sup>£</sup> | 88.92 (31.47)<sup>£</sup> |
| **Blood Cell Counts** (mean (SD)) |  |  |  |  |
| Platelets (K/μL) | 242.3 (70.4) | 241.7 (72.9) | 244.14 (67.71)<sup>£</sup> | 260.70 (71.18)<sup>£</sup> |
| WBC (K/μL) | 7.6 (2.1)* | 6.6 (2.2)* | 7.23 (2.16)<sup>‡</sup> | 6.57 (2.12)<sup>‡</sup> |

\*P-values comparison of the two sexes within Study cohort using a chi square test for categorical data, 2-sample t-tests for normally distributed continuous variables and Wilcoxon Rank Sums tests for non-normal continuous variables.
NHW: non-Hispanic White; AA: African American; PRISm: Preserved ratio impaired spirometry; FEV<sub>1</sub>: Forced expiratory volume in 1 second; FEV<sub>1</sub>/FVC: Ratio of the forced expiratory volume in one second to the forced vital capacity.
\*Significantly different $p \leq 0.01$ between races in COPDGene
<sup>‡</sup>Significantly different $p \leq 0.01$ between races in SPIROMICS

### COPD Phenotypes and Exposures

COPD was defined by spirometric evidence of airflow obstruction, which was computed as a ratio of post-bronchodilator forced expiratory volume at one second (FEV_1_) to forced vital capacity (FVC). The Global Obstructive Lung Disease (GOLD) system is used to grade COPD: in our smoking groups (current and former) GOLD 0 represents an individual without COPD (FEV_1_ > 80 %; FEV_1_/FVC ≥ 0.7), GOLD 1 (FEV_1_ ≥ 80 %; FEV_1_/FVC < 0.7), GOLD 2 (50% ≤ FEV_1_ < 80%; FEV_1_/FVC < 0.7), GOLD 3 (30% ≤FEV_1_ < 50%; FEV_1_/FVC < 0.7), and GOLD 4 (FEV_1_ < 30%; FEV_1_/FVC < 0.7), respectively represent the smoker control, mild, moderate, severe, and very severe stages of COPD. Individuals with an FEV_1_/FVC ≥ 0.70 and FEV_1_ % predicted ≤ 80% were defined as having Preserved Ratio Impaired Spirometry (PRISm) [37]. We use FEV_1_ as measured in liters as opposed to the race-based percent predicted which can create bias, but adjust for other covariates described below. Emphysema was captured as the log-transformed percentage of lung voxels with Hounsfield Units (HU)□<□−950 (%LAA950) on chest CT scan. This metric is also called percentage of low attenuation areas (%LAA). Current smoking status was defined as “former smokers” if they had not smoked any cigarettes within the last 30 days or “current smokers” if they had. Data to calculate the number of pack-years a person smoked were self-reported and calculated based on the packs of cigarettes smoked per-day multiplied by the total number of years smoked.

### Matched Non-Hispanic White and African American Race Groups

COPDGene non-Hispanic White (NHW) and African American (AA) groups had different sample sizes as well as key characteristics such as age, current smoking status, sex, and severity of COPD (GOLD Stage). Therefore, we applied a matching approach using SAS version 9.4 *SAS/STAT version 15.1, surveyselect* procedure to better match groups on these variables, with a particular focus on current smoking and GOLD stage. Details are provided in **Supplementary Methods.**

### Proteomic Platforms and Final Data Sets

Plasma protein levels were quantified with SomaScan and quality controlled by SomaLogic (Boulder, Co) [38]. Further details on SomaScan platforms are provided in **Supplementary Methods**. For COPDGene, the final matched race groups were 1,660 NHW and 1,513 AAs (**Table 1, Supplementary Methods**). For SPIROMICS, the final replication group was 1,792 subjects (1,459 NHW and 333 AA) (**Table 1)**.

### Covariate Adjustment

To account for potential confounding effects, we adjusted proteomic data for sex, age, and clinical center. Specifically, we fit an ordinary least squared regression model for each protein such that its abundance was used as the response variable and the three variables (sex, age, and clinical center) as covariates. The resulting residuals were used as input for downstream analysis.

### Network Analysis

*Network construction:* We used SmCCNet [30] to generate protein subnetworks associated with each COPD phenotype (FEV_1_ and %LAA950) and smoking (**Supplement Figure 3**). SmCCNet was originally developed to consider multiple omics data sets, so we modified the SmCCNet algorithm to a single omics setting by removing scaling between pairs of omics data. This proposed method has two implementations: one for continuous outcomes (applied to %LAA950 and FEV_1_) and one for binary outcomes (applied to smoking status). The continuous outcome scenario follows the SmCCA framework and implements sparse canonical correlation analysis. The binary exposure scenario implements sparse partial least square discriminant analysis (SPLS-DA) [39, 40], by performing a classification task under a supervised setting with a two-stage procedure. For the first step, the projection matrix is extracted with regular partial least square assuming a continuous phenotype. For the second step, the projected data is used to fit a logistic regression model. Details are provided in **Supplementary Methods**.

*Network trimming and summarization:* The subnetworks obtained through hierarchical clustering may still contain some proteins which are not strongly associated with the phenotype of interest. Therefore, our next step was to further trim the subnetworks such that only the most informative proteins were retained using the PageRank algorithm [41]. We then summarized each subnetwork using the NetSHy approach which applied principal component analysis (PCA) on the combination of both protein abundance and topological properties to obtain the first three low-dimensional summarization scores, referred to as NetSHy scores [42]. In all but one case noted in the Results, the top three scores accounted for over 40% of the cumulative variance explained. We calculated the correlation between each NetSHy score with the corresponding phenotype. Recall that each NetSHy score is a weighted average abundance of all proteins in the network with the relative weights determined by the corresponding loadings. By ranking absolute values of the loadings, we can identify top five proteins that contribute the most to each NetSHy score in each network. We denote these as <u>top five loading proteins</u>. We use the L2-norm explained, defined as the sum of squares of the top 5 protein’s loadings from each NetSHy PC, to check the total contribution of these proteins to their corresponding NetSHy PC. We found that among all 18 NetSHy PCs (6 networks X 3 PCs), 15 of them have at least 90% of the L2-norm explained, and all of them have at least 65% L2-norm explained by the top 5 proteins.

Based on the topology of each network, we compute the <u>total connection strength</u> of each protein by adding up all the edges connecting that protein to every other protein in the network. We define <u>hub proteins</u> as those proteins that have the top five largest total connection strength values (in some cases there are ties, see **Supplementary Table 3**). We use a ranking approach, as opposed to absolute cutoffs for the number of connections, as the density of the networks may vary.

*Statistical test for comparing subnetworks:* We quantify the similarities and differences between subnetworks associated with each phenotype and exposure across the two race groups using the p-norm difference test (PND) with the exponent p = 6, referred to as PND6, which was shown to be a top performing test by Arbet et al. [43]. For each phenotype and exposure, we compute a PND6 statistic which aggregates all the edge-wise differences across the two group-specific subnetwork adjacency matrices. Using a non-parametric permutation method, we derive the sampling distribution under the null hypothesis to generate the corresponding p-values. In our setup, p-values that are smaller than a significance level α correspond to rejecting the null hypothesis at the α level, indicating that the two comparing subnetworks are different. More details are provided in **Supplementary Methods**.

*Network projection:* In addition to a direct subnetwork comparison using the PND6 test statistic, we also investigate the similarities and differences between race-specific subnetworks by projecting a subnetwork derived from one race group onto another and vice versa. Specifically, we impose the subnetwork connectivity from one group onto the proteomic data of the other group to compute NetSHy scores as in [42], referred to as projection scores. We calculate correlations between these scores with each respective phenotype or exposure to statistically compare with the original correlations. This procedure is also used to compare subnetworks between COPDGene and SPIROMICS cohorts. Details are provided in **Supplementary Methods**.

### Network Quantitative Trait Loci (nQTL) Analysis

COPDGene WGS data was generated by the NHLBI Trans-Omics for Precision Medicine (TOPMed) program [44]. Details are provided in **Supplementary Methods**. For each subnetwork, we performed a genome-wide network quantitative trait locus (nQTL) analysis of the 3 inverse-normalized NetSHy scores (NetSHy1, NetSHy2, NetSHy3) assuming an additive model for genotype [42]. We regressed the NetSHy scores on each genetic variant separately adjusting for covariates depending on the phenotype used to generate the sub-network. For FEV_1_ and %LAA950 – the nQTL model was adjusted for sex, age, BMI, smoking status, and 6 genetic PCs to adjust for global ancestry . For smoking - the nQTL model was adjusted for sex, age, BMI, and 6 genetic PCs [45]. We conducted nQTL analysis on the University of Michigan Encore [46] server’s “Efficient and parallelizable association container toolbox” (EPACTS) [47]. Briefly, EPACTS efficiently performs statistical tests between phenotypes/exposure and sequence data through a user-friendly interface.

### Conditional nQTL Analysis

As a secondary analysis, we conducted genome-wide association tests for top proteins contributing to each NetSHy score, defined by their contribution to the NetSHy score. We regressed the inverse-normalized protein levels adjusting for covariates in the same manner as for the NetSHy network scores. If associations for phenotype and protein were observed in the same chromosomal locus, colocalization analysis was performed to assess whether the same genetic region contributed to both the genetic associations. If colocalization was observed, genome-wide analysis of phenotype was rerun with normalized protein value as an additional covariate, testing the hypothesis that the network quantitative trait loci (nQTLs) were driven by single protein quantitative trait loci (pQTLs). Further details are described in the **Supplementary Methods**.

### Pathway Overrepresentation Enrichment Meta-analysis

Proteins from each network were input into Metascape [48] as discrete lists. Uniprot identifiers were mapped to Entrez gene IDs. These genes were then assessed for enrichment in a variety of databases (Functional Set: Gene Ontology (GO): Molecular Functions; Pathway: GO: Biological Processes, Hallmark, Reactome, KEGG Pathway, WikiPathways, Canonical Pathways, BioCarta Gene Sets, PANTHER Pathway; Structural Complex: GO: Cell Components, CORUM). All proteins assayed by the SomaScan v4.0 platform were included as a background list for enrichment. Protein-protein interaction (PPI) networks obtained from STRING [49], BioGrid [50], OmniPath [51], and InWeb_IM [52] were additionally seeded with these genes and the MCODE algorithm [53] was used to identify subnetworks of connected proteins.

## Results

Despite matching some differences between NHW and AA still exist in the matching variable, but these differences are not clinically large. The biggest differences seen are with COPD Gold stages with AA having a larger percentage with normal lung function and a lower median number of pack-years of smoking. In the SPIROMICS cohort, which was not matched there are large differences in age, sex, smoking status, and severity of COPD. Both cohort’s AA population had higher levels of emphysema (**Table 1**). While the two cohorts are COPD cohorts, their recruitment criteria were different, and therefore there are difference in their overall characteristics with SPIROMICS being on average older, with a higher percentage of NHW, males, current smokers with a higher number of pack-years, more severe COPD and emphysema (**Supplement Table 2**).

### Protein Networks Associated with COPD Phenotypes and Smoking Exposure

#### Smoking

The NHW smoking network consisted of 34 proteins while the AA smoking network consisted of 17 proteins (**Figure 1**). Of those network proteins, only 27 and 7 proteins for NHW and AA respectively were significant in the univariate analysis at FDR < 0.10 (**Table 2, Supplement Table 3**). Across the two race groups, there were seven overlapping proteins including UCRP, PAP1, LPLC1, IGFBP-1, alkaline phosphatase placental type (ALPP), leptin, and EDIL3. In the NHW network, correlation between each protein and smoking status ranged from -0.20 to 0.36. The range of correlation between the proteomic data and smoking status was smaller in the AA network (-0.17 to 0.23). Correlations between networks in NHW and AA groups with the smoking exposure were 0.33 and 0.23, respectively (**Table 2**). Both networks displayed high connectivity such that each node was connected to every other node, leading to the corresponding network density equal to one. In both networks, ALPP had many heavily weighted connections. In particular, the connection strengths from ALPP to leptin, CRLD2, and GKN2 were 1, 0.79, and 0.76, respectively, in the NHW network. Similarly, in the AA network, ALPP was strongly connected to EDIL3 (1.0), leptin (0.9), and IGFBP-1 (0.67). As expected, by intersecting the lists of hub proteins and top loading proteins, we observed that hub proteins generally contributed more to the network summary scores than other proteins across the two race groups. For instance, in the NHW network, hub proteins such as ALPP, leptin, and PPBN were also among those with the largest loadings. Similarly, hub nodes in the AA network including ALPP, leptin, and trypsin-2 also contributed the most to the network summary score.

**Figure 1:**
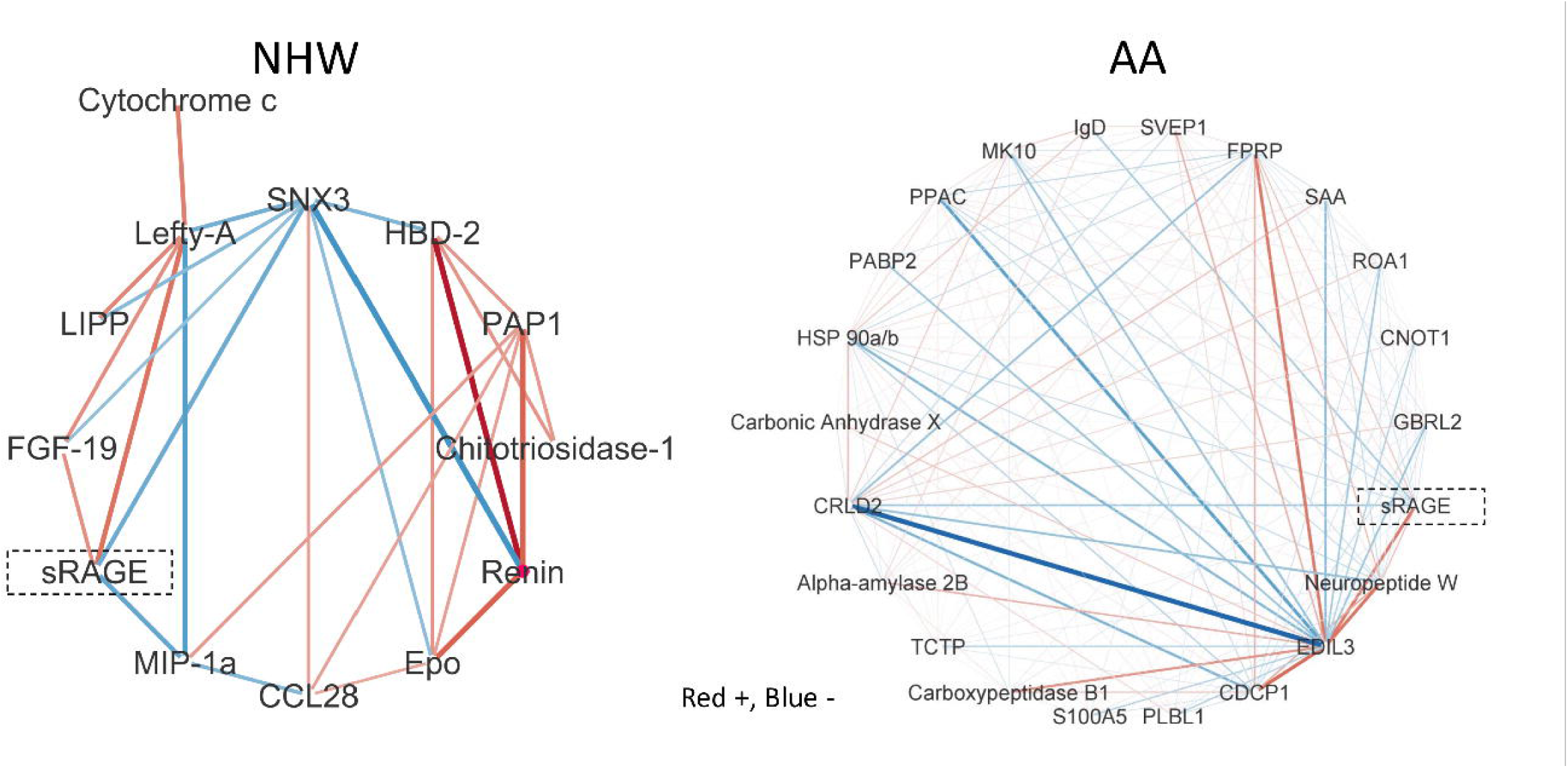
Networks associated with smoking exposure for NHW and AA populations. NHW network consists of 34 proteins while AA network has 17 proteins. Red edges connect proteins that have positive correlations with smoking exposure while blue edges link negatively correlated proteins. The line width is proportional to the connection strength. Correlations between networks in NHW and AA populations with the smoking exposure are 0.33 and 0.23, respectively.

**Table 2.** Summary results for each phenotype- and population-specific subnetwork. M denotes the number of proteins in each subnetwork. The number of proteins within the network that are significant in a univariate analysis are included (FDR <0.10). Correlations for the network are calculated by correlating the first three NetSHy scores and the respective phenotype.

| Phenotype | Population | M | Univariate<br>(FDR < 0.10) | Correlation between NetSHy scores and phenotype |  |  |
| --- | --- | --- | --- | --- | --- | --- |
|  |  |  |  | 1 | 2 | 3 |
| Smoking | NHW | 34 | 27 | 0.33 | 0.21 | 0.02 |
|  | AA | 17 | 7 | 0.23 | 0.14 | 0.03 |
| FEV<sub>1</sub> | NHW | 13 | 2 | 0.14 | 0.15 | 0.07 |
|  | AA | 22 | 1 | 0.13 | 0.09 | 0.01 |
| %LAA950 | NHW | 21 | 4 | 0.14 | 0.09 | 0.01 |
|  | AA | 104 | 6 | 0.12 | 0.09 | 0.12 |

We used a statistical approach to compare the adjacency matrices representing the two race-specific networks. Given that the two networks had different sizes (34 vs. 17 proteins), we found a union set of 44 proteins present in either or both networks, prior to calculating the p-norm difference test with exponent equal to 6 (PND6) (See Methods). **Table 3** shows the resulting test statistics and p-values when comparing smoking-associated networks to indicate that networks associated with smoking are similar across NHW and AA race groups (PND6 = 0.340, p-value = 0.955). **Supplement Figure 4a** displays the corresponding heatmap for edge-wise differences in networks associated with smoking exposure between NHW and AA groups. In alignment with the PND6 test, we observed more white or lighter red areas, highlighting the similarity of smoking-associated networks across the two race groups. Additionally, **Table 4** summarizes the similarities and differences between smoking-associated subnetworks by projecting a subnetwork across race groups and/or cohorts, which is a complementary approach that does not require adjacency matrices in each group. Within the COPDGene, we computed the cross-race correlations by projecting the NHW subnetwork onto AA data and vice versa, and we observed similar correlations across the two race groups. Specifically, when the AA subnetwork (C-AA) was projected to NHW proteomic (C-NHW) data, the first two projection correlations were 0.354 and 0.125, respectively. The original correlations, 0.329 and 0.208, fell within the corresponding 95% bootstrap confidence intervals (CIs) of (0.303, 0.403) and (0.057, 0.202), respectively. Similarly, when we projected the NHW subnetwork (C-NHW) onto AA (C- AA) data, the corresponding 95% CIs also captured the observed correlations, demonstrating the similarity between subnetworks across the two race groups within the same cohort.

**Table 3:** Results of direct network comparison and projection using subnetworks associated with smoking exposure, FEV_1_, and %LAA950 across NHW and AA. M denotes the number of proteins in each network. To perform the direct network comparison, we used the p-norm difference test (PND) test with the exponent p = 6, referred to as PND6 (Arbet et al., 2021). Small p-value indicates that two networks being compared are different. NetSHy correlations are the correlations between the first three NetSHy scores and each respective phenotype and exposure, while projection correlations are calculated using the projection scores.

| Across cohorts | C-NHW | S-NHW | **0.373**<br>(0.334, 0.416) | **0.186**<br>(0.135, 0.235) | **0.027**<br>(0.002, 0.082) |
| --- | --- | --- | --- | --- | --- |
| Across populations & cohorts | C-AA | S-NHW | **0.393**<br>(0.347, 0.432) | 0.031<br>(0.002, 0.130) | **0.011**<br>(0.001, 0.059) |
| AA Data |  |  |  |  |  |
|  | Network | Data | Component |  |  |
|  |  |  | 1 | 2 | 3 |
| Original | C-AA | C-AA | 0.235 | 0.143 | 0.031 |
| Across populations | C-NHW | C-AA | **0.232**<br>(0.188, 0.272) | **0.126**<br>(0.082, 0.178) | **0.020**<br>(0.001, 0.057) |
| Across cohorts | C-AA | S-AA | **0.249**<br>(0.163, 0.325) | **0.073**<br>(0.004, 0.199) | **0.019**<br>(0.003, 0.149) |
| Across populations & cohorts | C-NHW | S-AA | **0.245**<br>(0.175, 0.317) | **0.116**<br>(0.008, 0.229) | **0.019**<br>(0.002, 0.130) |

**Table 4:**
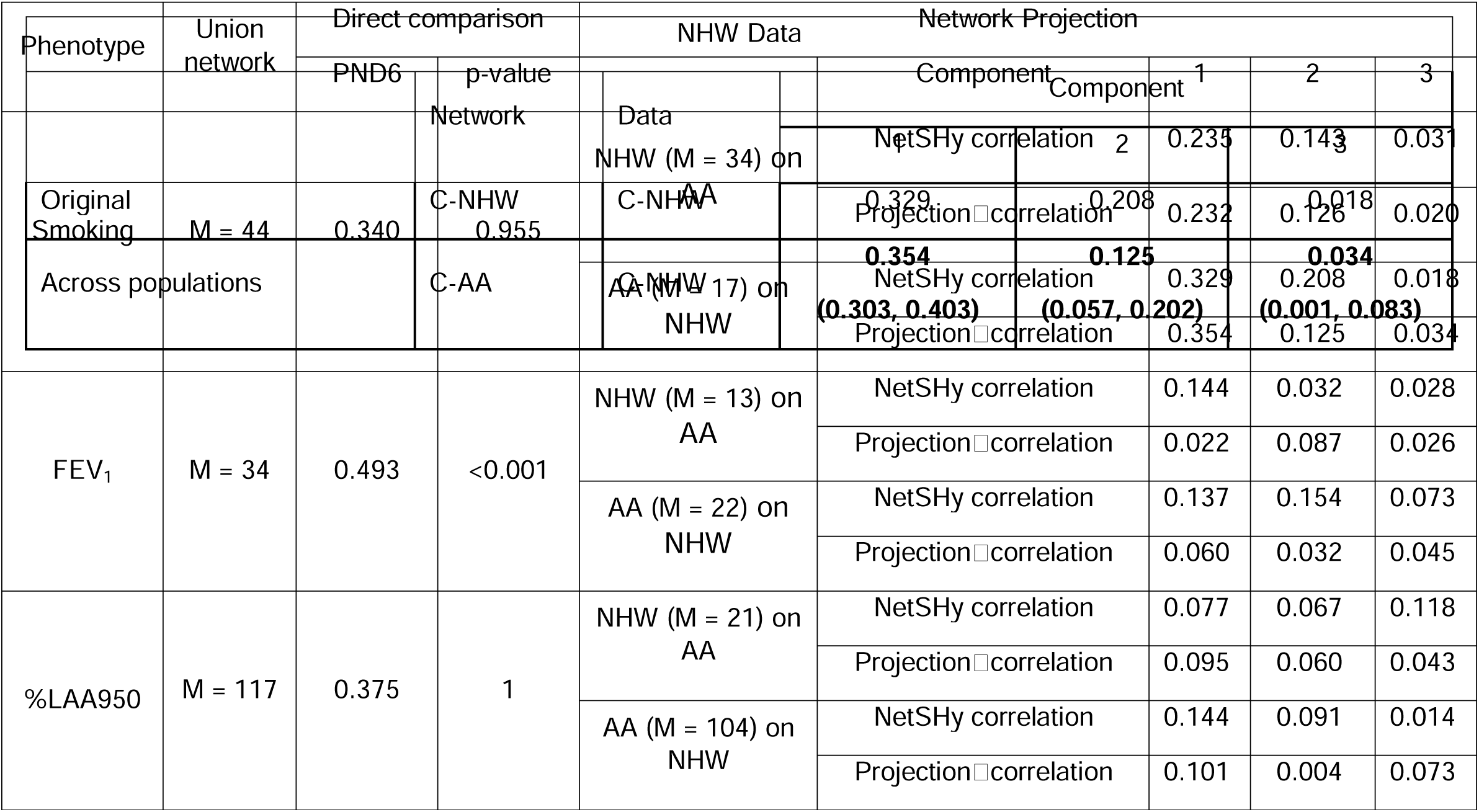
Summary results of network projections using subnetworks associated with smoking exposure across and AA, in two independent cohort studies COPDGene (C) and SPIROMICS (S)

**Table 5.** Results of genome-wide significant association tests of NetSHy scores. The genetic association with the smallest p-value for each network is listed, along with its minor allele frequency (MAF), effect size (Beta), and the total number of SNPs associated (No. Hits). Colocalization tests were performed to test whether the sub-networks share genetic signals with any of the top five proteins contributing the most to it. A posterior probability (PP) of the shared variant hypothesis (H4) greater than 0.9 indicates probable colocalization of genetic signals. For colocalized signals, we further ran sensitivity analyses that used the same genetic association regression model as previously but adjusting for the levels of the protein. Significant results (*p* ≤ 5x10^-8^) are highlighted in bold. AA: African American. NHW: Non-Hispanic white.

| Phenotype | Population | NetSHy Score | Variant (rsID) | MAF | Beta | p-value | No. Hits | Closest Gene | Annotation | Colocalization with pQTL | PP (H4) | p-value post sensitivity |
| --- | --- | --- | --- | --- | --- | --- | --- | --- | --- | --- | --- | --- |
| %LAA950 | AA | 2 | chr2:134214910 (rs72846742) | 0.083 | 0.390 | **1.0E-08** | 1 | *MGAT5* | Intronic | None | - | - |
|  | NHW | 2 | chr9:133262254 (rs8176693) | 0.080 | -0.392 | **7.2E-09** | 9 | *ABO* | Intronic | None | - | - |
|  |  | 3 | chr9:133273983 (rs992108547) | 0.196 | 0.287 | **1.2E-10** | 17 | *ABO* | Intronic | Cadherin 17 | 99.50% | **3.30E-03** |
|  |  |  | chr19:51127225 (rs2258983) | 0.422 | -0.217 | **2.6E-09** | 3 | *SIGLEC9* | Nonsynonymous coding | Cadherin 17 | 0.0075% | - |
| FEV<sub>1</sub> | NHW | 2 | chr1:225890637 (rs360060) | 0.325 | 0.358 | **2.2E-22** | 65 | *LEFTY1* + *AL117348.2* | Intronic | Left right determination factor 2 | 99.50% | **3.51E-04** |
| Smoking | AA | 1 | chr2:232409765 (rs56080708) | 0.0708 | -0.446 | **7.1E-10** | 3 | *ALPG* | Nonsynonymous coding | Alkaline phosphatase placental type | 99.90% | **2.34E-01** |
|  | NHW | 1 | chr2:232421944 (rs12478529) | 0.237 | -0.418 | **9.4E-24** | 74 | *ALPG* | None | Alkaline phosphatase placental type | 96.70% | **2.86E-02** |

We further projected the subnetworks derived from COPDGene (C) onto the data in SPIROMICS (S) to assess the replicability of the subnetworks across independent cohorts. By projecting the NHW subnetwork derived from COPDGene (C-NHW), onto the NHW data in SPIROMICS (S-NHW) we obtained the first two cross-cohort correlations of 0.373 and 0.186, respectively. Note that the 95% CI of the first projection component (0.334, 0.416) was significantly higher than the original correlation of 0.329. Similarly, when we projected the COPDGene AA subnetwork (C-AA) onto SPIROMICS NHW (S-NHW) data, the first projection correlation was 0.393 and its 95% CI was (0.347, 0.432). Once again, the confidence interval was higher than the original correlation of 0.329. Such consistent projection correlations indicate a high level of replicability of the subnetworks associated with smoking exposure across independent cohorts. In a similar manner, we projected the C-AA subnetwork onto the SPIROMICS AA (S-AA) data and also observed similar results (**Table 4**). In summary, these results provide further evidence of the replicability of the smoking subnetworks across cohorts, even when considering different race groups.

##### FEV_1_

There were 13 and 22 proteins present in the NHW and AA networks for FEV_1_, respectively, with sRAGE present in both networks (**Figure 2**). Of those network proteins, only 2 and 1 protein(s) for NHW and AA respectively were significant in the univariate analysis at FDR < 0.10 (**Table 2, Supplement Table 3**). In the AA network, sRAGE was strongly connected to Carboxypeptidase B (1.0) and EDIL3 (0.53) while displaying relatively weaker relationships (< 0.33) with the remaining nodes. In the NHW network, sRAGE showed strong connections to Renin (0.93) and Lefty-A (0.7) while maintaining moderate relationships of at least 0.5 to other proteins. Correlations between individual proteins with FEV_1_ ranged from -0.11 to 0.1 in the NHW network and from -0.09 to 0.12 in the AA network. Correlations between NetSHy1 of networks derived from NHW and AA participants with FEV_1_ were 0.13 and 0.14, respectively (**Table 2**).

**Figure 2:**
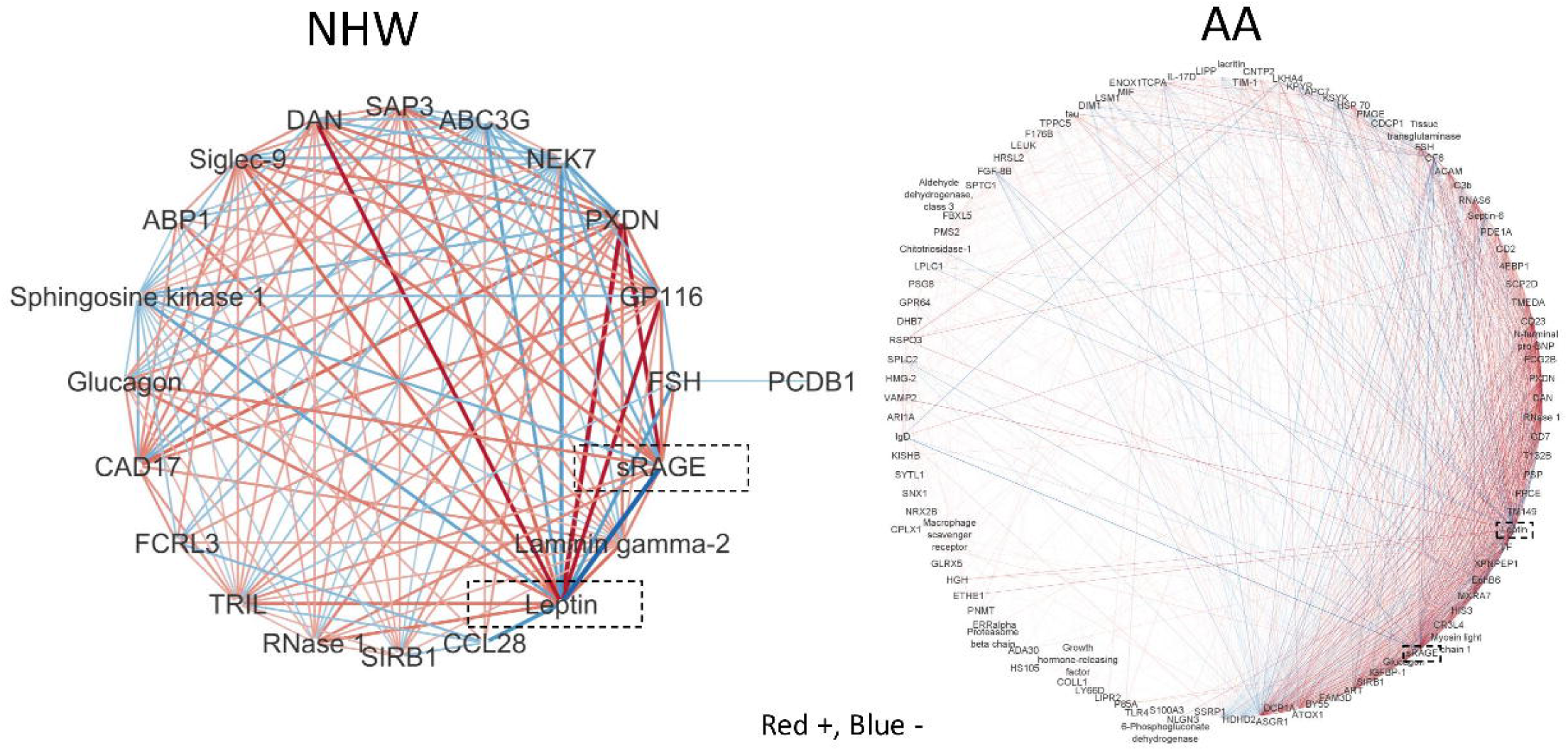
Networks associated with FEV1 for NHW and AA populations. NHW network consists of 13 proteins while AA network has 22 proteins. Red edges connect proteins that have positive correlations with smoking exposure while blue edges link negatively correlated proteins. The line width is proportional to the connection strength. Correlations between networks in NHW and AA populations with the smoking exposure are 0.13 and 0.14, respectively.

We next investigated potential overlap between the NHW and AA networks. Using the PND6 method, we found a significant difference between the two networks (p-value < 0.001, **Table 3, Supplement Figure 4b**). The projection approach also showed *poor performance*, suggesting notable differences between the FEV_1_ networks across the two race groups. We further projected the subnetworks derived from COPDGene (C) onto the data in SPIROMICS (S) to assess the replicability of the subnetworks across independent cohorts. By projecting the C-NHW subnetwork onto the S-NHW data and vice versa, we found that the corresponding 95% CIs also captured the original correlations, suggesting some degree of replicability across cohorts for the same race group (**Supplement Table 3a**). However, the CIs were relatively wider than with smoking, which might be due to more variation in the subnetworks associated with this phenotype. These observations indicate some moderate degree of transferability of FEV_1_ associated networks across cohorts for the same race group. However, the results also highlight potential variations in the subnetworks associated with FEV_1_ across race groups, emphasizing the importance of considering group-specific characteristics when studying this phenotype.

### %LAA950

There were 21 and 104 proteins present in NHW and AA networks for %LAA950, respectively (**Figure 3**). Of those network proteins, only 4 and 6 proteins for NHW and AA respectively were significant in the univariate analysis at FDR < 0.10 ((**Table 2, Supplement Table 3**). The AA network is notably larger and denser, and was the only network where the top 3 summarization scores explained less than 40% of the variability (23% variability explained). Despite this difference there were many consistencies. The two networks had seven proteins in common: PXDN, DAN, FSH, sRAGE, Glucagon, SIRB1, RNase 1, and Leptin. In the NHW network, the range of correlations between each protein and %LAA950 was between -0.12 and 0.09, which was similar to that in the AA race group. Correlations between networks derived from NHW and AA groups with %LAA950 were 0.14 and 0.12, respectively (**Table 2**). Like smoking, the two networks associated with %LAA950 are similar across NHW and AA groups (**Table 3, Supplement Figure 4c**). This was also consistent with the projection analysis (**Supplement Table 3b**) where we found notable similarities between subnetworks associated with %LAA950 across the two race groups within the same cohorts. Furthermore, when comparing the subnetworks associated with %LAA950 across independent cohorts, we also observed consistency in the projections (**Supplement Table 3b**).

**Figure 3:**
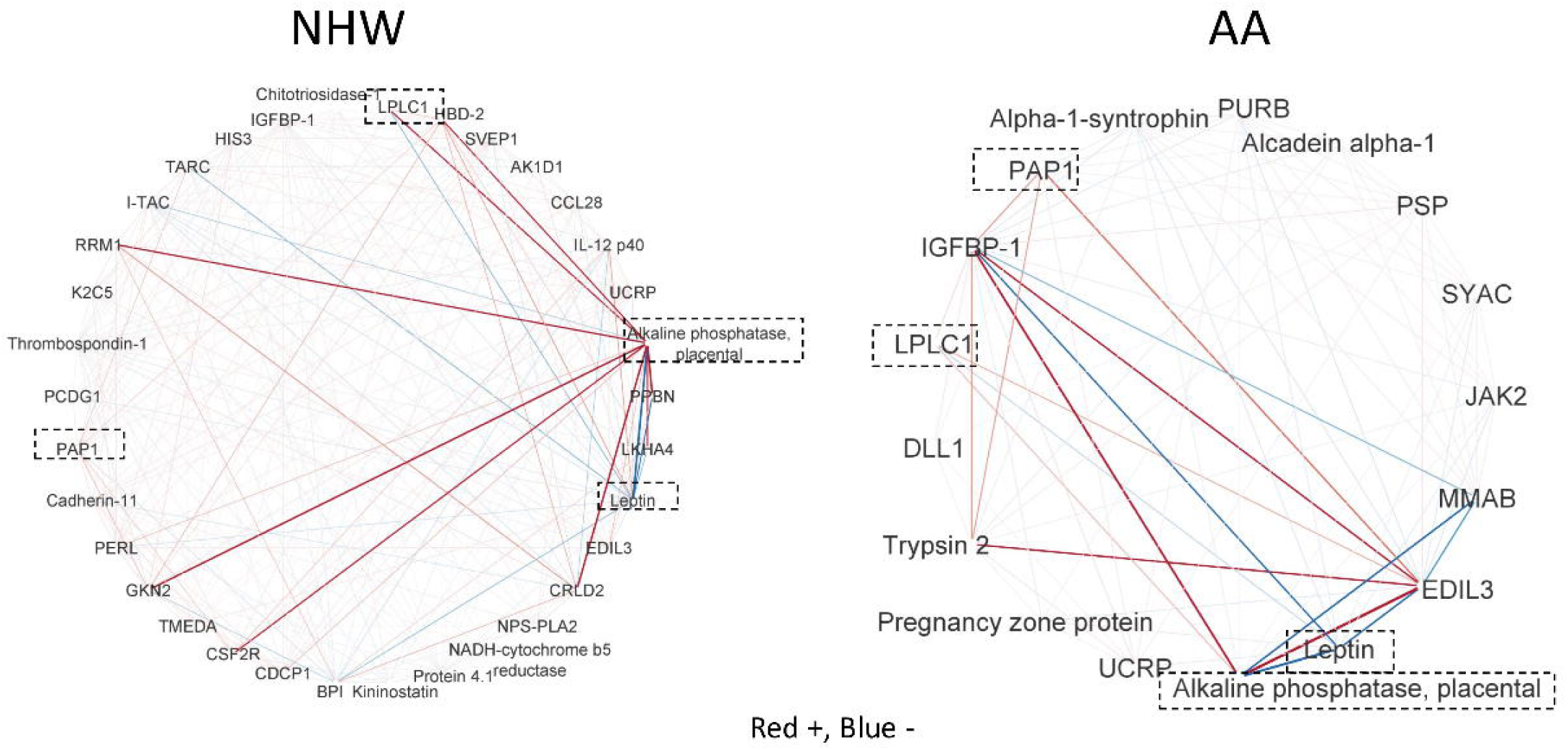
Networks associated with %LAA950 for NHW and AA populations. NHW network consists of 21 proteins while AA network has 104 proteins. Red edges connect proteins that have positive correlations with smoking exposure while blue edges link negatively correlated proteins. The line width is proportional to the connection strength. Correlations between networks in NHW and AA populations with the smoking exposure are 0.14 and 0.12, respectively.

#### Enrichment

We performed enrichment of individual proteins within networks and meta-analysis across networks through MetaScape. Significantly enriched pathways are shown in **Figure 4**. Top shared pathways identified through meta-analysis include response to hormone (enriched in all gene lists), and regulation of cell activation and response to bacterium enriched in five gene lists **(Figure 4a).** Many additional pathways were enriched in multiple gene lists in meta-analysis. Individual enrichment analysis also showed gene lists were enriched for many disease-relevant pathways. For example, in addition to observing many proteins in networks associated with inflammatory and antimicrobial processes, we observe VEGFA-VEGFR2 signaling enriched in %LAA950 NHW, FEV_1_ NHW, and FEV_1_ AA networks (**Figure 4b**).

**Figure 4.**
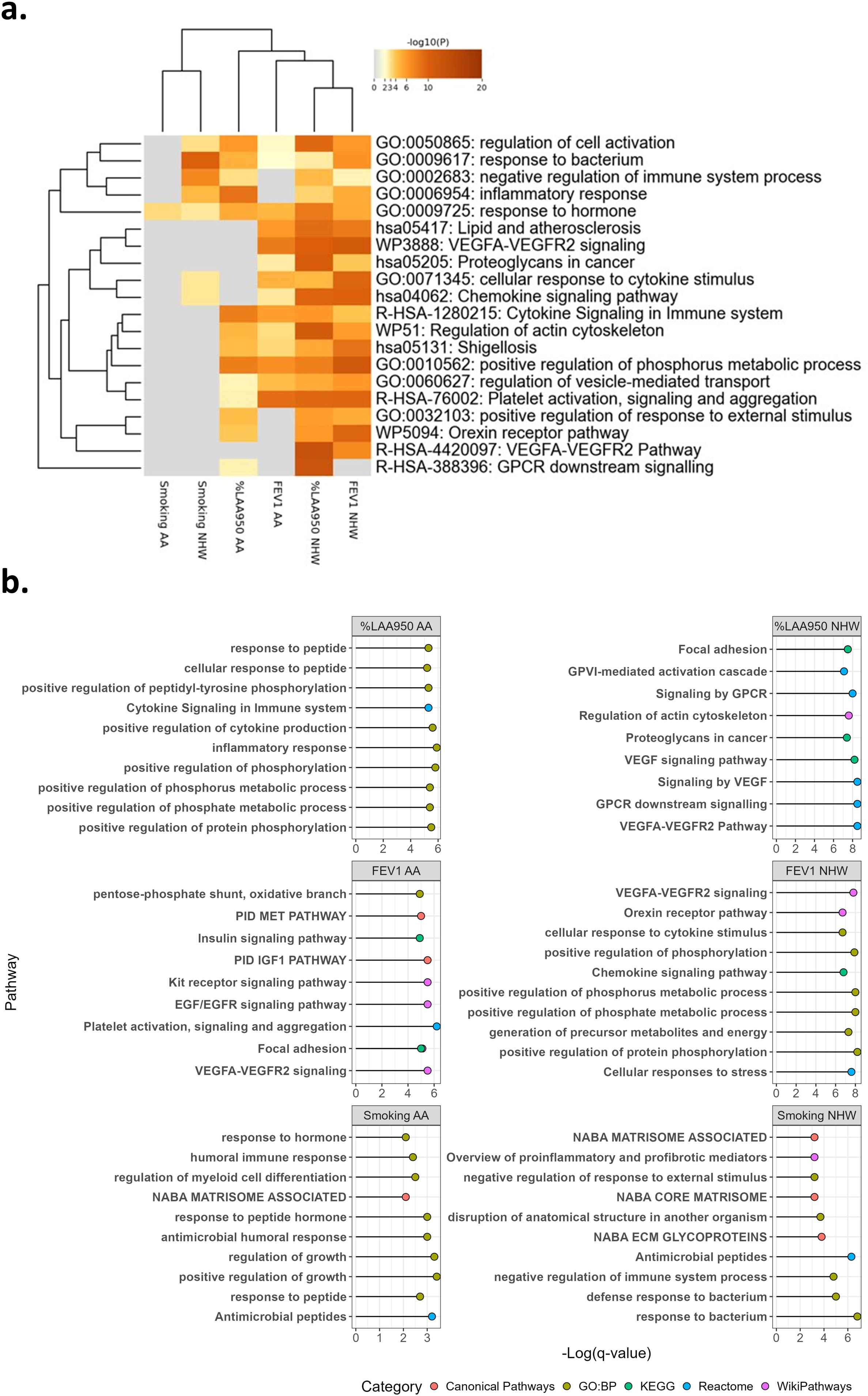
Pathway Enrichment Analysis. A. Heatmap of top 20 significantly enriched pathways across protein lists identified through Metascape meta-analysis. Enriched pathways are colored by –log10(p-value). B. Top 10 enriched pathways by network.

### Network QTLs (nQTLs) Show Genetic Underpinnings of COPD Protein Networks

We tested for association between the top 3 NetSHy scores of each protein network and common genetic variants from WGS. Seven NetSHy scores were associated with at least one variant at a genome wide significant level (**Table 5**, **Figure 5**). NetSHy1 of smoking in both AA and NHW participants show genetic association signals on 2q37.1 within or near the gene *ALPG*. NetSHy2 of FEV_1_ in NHW participants is associated with variants on chr1 near *LEFTY1*. NetSHy2 of %LAA950 in AA participants is associated with a single variant in *MGAT5*, and NetSHy2 and NetSHy3 of %LAA950 in NHW participants show associations with the *ABO* locus. NetSHy3 of %LAA950 in NHW additionally shows an association signal on chr19 within the gene *SIGLEC9*. Both *ABO* lead variants have previously been found to be associated with lung function. Rs8176693 was nominally associated with FEV_1_/FVC in a European population [54] and rs9921085 is associated with both FEV_1_ *(p*-value = 1.00 x 10^-14^) and FVC *(p*-value = 1.10 x 10 ^-14^) in the UK Biobank [55].

**Figure 5.**
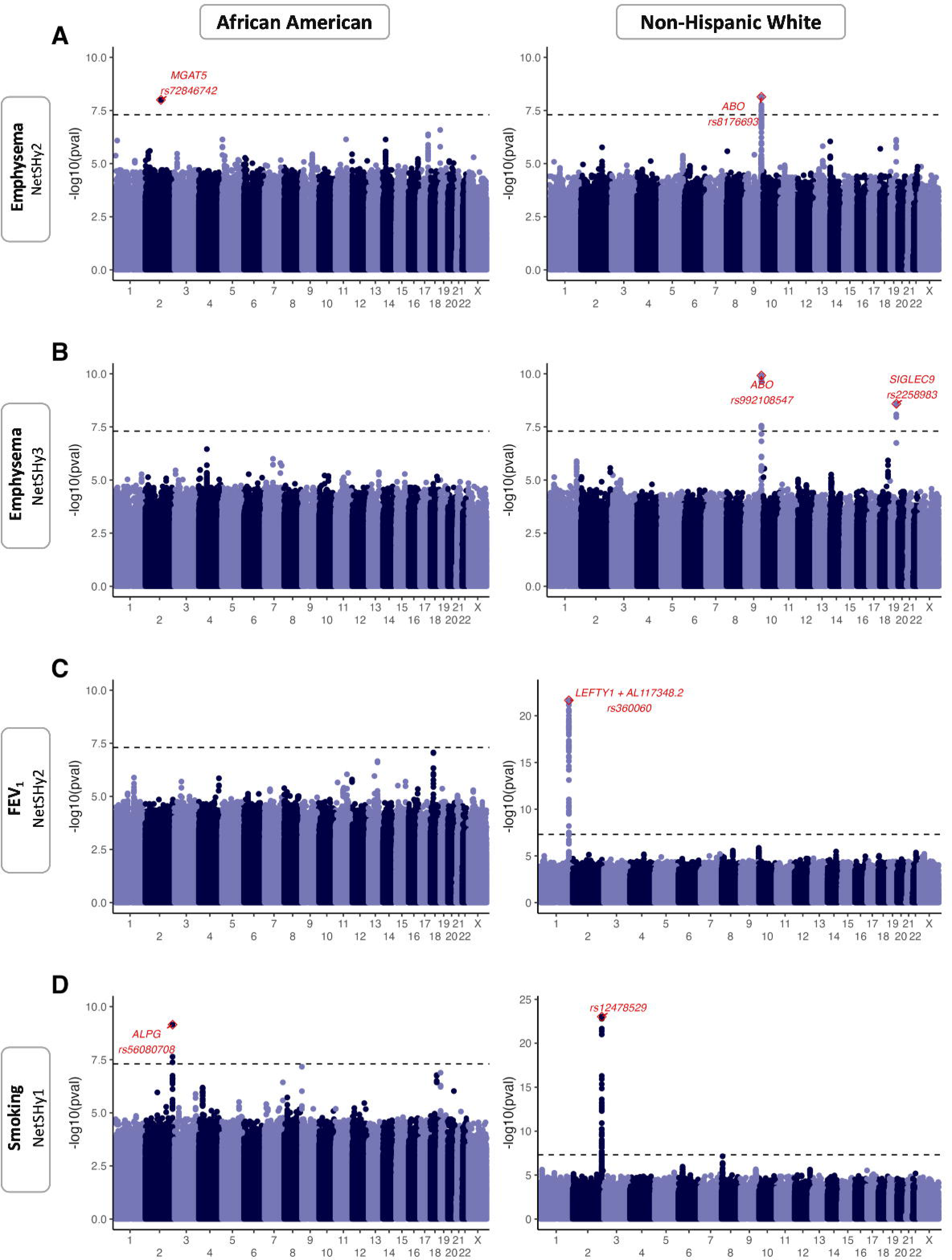
Network QTL Associations with Protein Network NetSHy Summary Scores. Manhattan plots display the significance (-log10 of p-values, y axis) of genome-wide association tests across all chromosomes (x axis) in African Americans (left panels A-D), and non-Hispanic whites (right panels, A-D). Each row corresponds to the following associations: A) Emphysema NetSHy2, B) Emphysema NetSHy3, c) FEV1 NetSHy2, D) Smoking NetSHy1. The top SNP of each association signal is highlighted in red, and it is labeled with the rsID of the SNP, along with the name of the closest gene, when applicable.

We next assessed whether these genetic associations were driven by top proteins in networks. For each NetSHy score with a significant association, we ran a genome-wide association scan for the top 5 loading proteins contributing to each NetSHy score. We identified associations with proteins in %LAA950 NHW NetSHy 2 (Ganglioside GM2 activator), %LAA950 NHW NetSHy3 (Cadherin 17 and sRAGE), FEV_1_ NHW NetSHy2 (Regenerating islet derived protein 3 alpha), Smoking AA NetSHy1 (Cob(I)yrinic acid a,c-diamide adenosyltransferase mitochondrial, alkaline phosphatase placental type, and insulin growth factor binding protein 1), and Smoking NHW NetShy1 (Gastrokine 2, Interleukin 12 subunit beta, Alkaline phosphatase placental type, and alkaline phosphatase placental like 1) (**Supplement Tables 4**).

In each instance where an nQTL and a single-protein genetic association were on the same chromosome, we tested for colocalization of these signals using *coloc*. When single protein and nQTL signals colocalized, we reran the associated GWAS with the single protein abundance values included as a covariate to serve as a conditional analysis. After conditional analysis, 4 NetSHy associations with genetic loci of the 7 remain: NHW %LAA950 NetSHy3 –*SIGLEC9*, AA %LAA950 NetSHy2 - *MGAT5*, NHW %LAA950 NetSHy2 – *ABO*, and NHW FEV_1_ NetSHy2 – *LEFTY1* (**Supplement Table 4**).

### Discussion

#### Summary

We used SmCCNet to generate protein correlation networks associated with FEV_1_,

%LAA950, and smoking status separately in NHW and AA COPDgene participants, containing 13 to 104 proteins. We used smoking exposure as a paradigm to develop methods and contrast race groups as smoking has been well studied. We then used the same approach to investigate other COPD phenotypes such as FEV1 and %LAA, where our understanding was comparatively limited. The derived networks demonstrated stronger or as strong correlations with phenotypes and exposure than individual proteins demonstrating the beneifts of a network approach. Smoking and %LAA950 networks were similar between NHW and AA, and replicated well in the SPIROMICS cohort, while FEV1 networks showed notable differences across the two groups and lower level of replicability.

We ran genome-wide association study analysis on NetSHy scores to identify potential genetic variants associated with the protein networks, which we refer to as nQTLs. Finally, we assessed whether discovered nQTLs were independent of genetic association signals of single top proteins included in the network and identified three genetic variants associated with %LAA950 networks. Through this work, we have identified novel networks of correlated proteins related to COPD phenotypes of interest, as well as common genetic variants associated with these networks. It is worth noting that at many of the proteins in the identified networks were not significantly correlated with the respective phenotype/exposure (**Table 2**). This demonstrates the advantages of a network approach, which enabled the identification of proteins that were not identified on their own but appear to play a supplementary role in influencing the outcome of interest through their interactions with other proteins that do have a strong association with the phenotype/exposure.

Enrichment analysis of networks demonstrates that network proteins across the phenotypes are associated with processes and pathways such as response to bacterium and antimicrobial peptides, hormone activity, extracellular matrix signaling, and interferon signaling. Antimicrobial proteins include UCRP and LPLC1 in our smoking networks, as well as proteins such as MIP1a and IgD in FEV_1_ networks, and PXDN and RNase1 in %LAA950 networks.

UCRP is integral to the response to infection of multiple respiratory pathogens, including influenza and SARS-CoV-2 [56, 57]. UCRP has previously been demonstrated to be upregulated at the RNA level in alveolar macrophages from COPD patients with more severe disease (based on GOLD staging) [58]. LPLC1 is thought to be involved in innate immune responses to bacterial infection, including in the lung [59]. LPLC1 has previously been demonstrated to be upregulated in sputum of smokers with and without COPD [60].

Furthermore, protein levels in sputum are correlated with smoking pack-years and spirometric measures of lung function (FEV_1_ & FEV_1_/FVC) [61]. MIP-1a is an inducible chemokine that promotes inflammation and monocyte and macrophage recruitment. Gene and protein expression is increased in COPD PBMCs relative to healthy controls [62] as well as in sputum [63]. MIP-1a has also been shown to promote tight junction injury in airway epithelium [62]. IgD is the major antigen receptor type on peripheral B-cells. It induces TNF, IL1B, and IL1RN, in addition to other cytokines [64]. Serum IgD has previously been shown to be increased in COPD subjects [65]. PXDN is a heme-containing peroxidase secreted into extracellular matrix that is involved in extracellular matrix formation. PXDN also directly binds gram-negative bacteria in innate immune response, contributing to lung host defense [66]. RNase 1 is an endonuclease targeting single- and double-stranded RNAs. RNASE1 has previously been seen to be upregulated at the gene expression level in PBMCs from COPD patients compared to those from healthy controls [67].

Networks also contain hormones and proteins involved in hormone signaling. These include leptin and IGFBP-1 in smoking networks, glucagon in %LAA950 networks, and renin in FEV1 networks. Leptin is an adipocyte-derived hormone with pro-inflammatory effects. There is conflicting evidence of altered leptin concentrations in COPD [68–70]. Low levels of IGFBP-1 which binds both IGF 1 and 2, can indicate impaired glucose tolerance, vascular disease, and hypertension. IGF and IGFBP concentrations have been shown to be altered in COPD and smoking [71]. Glucagon is a pancreatic hormone involved in glucose metabolism and homeostasis and has been shown to reduce airway hyperresponsiveness [72]. Renin is an endopeptidase secreted by the kidneys that targets angiotensinogen, resulting in elevated blood pressure and vasoconstriction [73]. Upregulation of renin-angiotensin signaling can drive pulmonary fibrosis [74]. Angiotensin II regulates response to lung injury and apoptosis in alveolar epithelium [75] and there is some evidence that angiotensin-converting enzyme inhibitors and related drugs result in reduced exacerbations and mortality in COPD [76, 77].

Networks additionally contain molecules involved in tissue remodeling in COPD [78]. For example, our FEV_1_ networks contain sRAGE, a soluble receptor that binds advanced glycosylation end products, which accumulate in vascular tissues during aging. COPD patients show lower plasma and serum levels of sRAGE. Additionally, sRAGE levels are associated with emphysema severity and reduced FEV_1_ [79]. Smoking networks contain molecules such as EDIL3 and CRLD2. EDIL3 (EGF-like repeat and discoidin I-like domain-containing protein 3) is an integrin ligand that promotes adhesion of endothelial cells and is involved in angiogenesis and vascular remodeling. Plasma levels of EDIL3 have been shown to be decreased in COPD patients and associated with increased risk of acute exacerbation [80]. CRLD2 (Cysteine-rich secretory protein LCCL domain-containing 2) [CRISPLD2] is a secreted protein that promotes matrix assembly and modulates airway branching and alveogenesis [81]. Glucocorticoid treatment increases gene and protein expression in airway smooth muscle cells, which in turn regulates cytokine levels [82]. Heterozygous knockout mice display features similar to bronchopulmonary dysplasia [83]. CRLD2 has also been shown to attenuate inflammatory signaling induced by LPS in lung fibroblasts and epithelial cells.

Note that some of the proteins above reached nominal significance (p<0.001) in a univariate analysis (**Supplement Table 3**) with the respective exposure/outcome but very few reached statistical significance accounting for multiple testing (FDR < 0.10). This further illustrates the benefits of a network approach for identifying proteins that may not have the strongest univariate signal but may have strong interactions with other proteins related to the exposure/outcome.

#### nQTL Findings

We identified 7 nQTL signals for 6 unique NetSHy scores. nQTLs may play a role in the regulation of the network as opposed to individual pQTL which may only affect a single protein. As nQTLs may be driven by a single strong pQTL, we examined pQTLs for top network proteins and performed colocalization analysis. Four of the 7 nQTLs remained associated after conditional analysis adjusting for protein levels of top network proteins with colocalized pQTL protein values. These signals are a variant (rs72846742) on chr2 with AA %LAA950 NetSHy2, a locus overlapping *SIGLEC9* on chr19 with NHW %EMP NetSHy3, variants in the *ABO* locus with NHW %LAA950 NetSHy 2, and a locus on chr1 with NHW FEV1 NetSHy2. We note that while SIGLEC9 was not one of the top 5 protein loadings for NHW %LAA950, it is present in the network. rs72846742 has been previously associated with smoking intensity [84] and is within the first intron of *MGAT5* (alpha-1,6-mannosylglycoprotein 6-beta-N-acetylglucosaminlytransferase). It has also been shown to be an eQTL for *MGAT5* in blood by the eQTLGen consortium [85]. This gene encodes a glycosyltransferase primarily implicated in cancer. A recent study reports *MGAT5* genetic variation associated with COPD in a Chinese population [86].

The *ABO* locus has been extensively studied and variants in this gene have been associated with increased risk of numerous diseases. Despite multiple studies of *ABO* allele frequencies in COPD, no consistent association with disease or related phenotypes has been reported. The lead variant, within an intron of *ABO*, has been shown to act as both an eQTL and pQTL for *ABO* [87, 88] and has been associated with numerous phenotypes generally related to blood traits and cardiovascular disease.

Genes within the chr1 locus associated with NHW FEV_1_ NetSHy2 include *EPHX1*, *TMEM63A*, *LEFTY1*, *LEFTY2*, and *PYCR2*. We note that *LEFTY2* encodes left-right determination factor 2, a protein that is within the NHW FEV_1_ network despite not being a top protein loading on NetSHy2. This protein is a secreted ligand that binds TGF-beta receptors. TGF-beta signaling has been implicated in many aspects of COPD [89]. The lead variant in the locus, rs360060, has been shown to act as an eQTL for TMEM63A, LEFTY1, and EPHX1, and is predicted to most likely affect TMEM63A by the OpenTargets Platform [88]. The variants on chr19 are proximal to or within the gene body of *SIGLEC9*. Protein levels of *SIGLEC9* have been shown to be increased in plasma and neutrophils from COPD patients [90] and one variant, rs2075803, has previously been associated with higher exacerbation frequency and greater emphysema in a small cohort [91]. As many nQTL signals seemed driven by single-protein associations, future applications of this framework may address this through approaches such as regressing pQTL signals from the protein data [92].

It is important to note that this work was performed using SomaScan platform data, and although there was replication in an independent cohort for the same platform, our findings may not replicate across other proteomic assays. Furthermore, although SomaScan is one of the most comprehensive proteomic profiling methods, it only captures a subset of the proteome so may be missing proteins in the network. However, our genetic investigation of the FEV_1_ networks showed signals in loci well-studied in the context of COPD, such as *EPHX1* [93, 94], although the EPHX1 protein was not included in the SomaScan panel. This finding suggests that the protein networks and its genetic associated loci can capture biologically meaningful signals involved in COPD, even if they are not directly assayed in our study. In the future as platforms become more comprehensive, we will be able to expand on these networks in addition to incorporating other omics measurements. In addition, our results are subject to sources of noise inherent in these types of studies including the use of blood, as opposed to primary tissue, non-fasting measurements, and differences in medication use.

Across all network results, the respective networks had at most 0.33 correlation with smoking and at most 0.14 with FEV_1_ or %LAA950. Although the correlations with the two phenotypes may not seem strong, they were still larger than the correlation values observed for individual proteins (maximum correlation found for any protein was 0.12 for both phenotypes and race groups) and consistent with what we have observed in our previous biomarker studies [17, 79].

We decided to analyze NHW and AA participants separately within COPDGene for a variety of reasons. In COPDGene, NHW and AA participants display major differences in terms of demographics and disease severity. We implemented a matching scheme to better match NHW and AA groups on age, GOLD stage, and smoking status. In spite of this, groups still exhibited some differences in demographics and disease. To further address demographic confounding with omics signals, we regressed age, sex, and clinical center from the proteomic data prior to network generation. We decided to only include non-modifiable covariates which are unlikely to be influenced by disease in our regression model. Additionally, matching allowed us to down-sample NHW participants to a sample size closer to the AA group, mitigating differences in results that may have been driven by power/sample size issues, which occurs in many studies where data sets from European race/ancestry are typically much larger than other groups. Finally, we assessed whether networks derived from one race group replicated in the other group in terms of both network structure and NetSHy scores. We found that the smoking and %LAA950 networks were replicated across the race groups indicating shared interactions, even when all proteins in the network did not overlap. On the other hand, FEV1 did not show strong replication across race groups and/or study cohorts. This is not surprising given that spirometry generally shows a great degree of variability [95, 96]. Consequently, networks associated with FEV1 may capture such inherent variability, potentially reducing their replicability.

Although there were many similarities, we emphasize that any observed differences between race groups are likely the result of biases in sampling and potentially driven by social determinants of health; differing results between race groups do not indicate nor support differing biology between these groups. SDoH may induce proteomic changes leading to increased inflammation [97]. When examining self-rated health (SRH) data, poor SRH scores are linked to a rise in inflammatory plasma proteins such as leptin in CVD populations.

Additionally, SDoH variables such as education i.e., university degree attainment, while typically associated with poorer health outcomes were not related to SRH [98]. This current work shows leptin’s inflammatory nature being implicated in COPD. There are few studies examining SDoH and COPD and pose a novel path forward for investigation.

## Conclusion

In this work, we constructed protein networks that are related to COPD-relevant phenotypes, namely FEV_1_ and %LAA950, and the primary exposure of smoking, separately in NHW and AA COPDGene participants. We demonstrate the ability to derive sparse protein networks associated with these phenotypes that replicate both across race sub-groups and across cohort studies. By leveraging NetSHy network summarization scores, we were further able to identify common genetic variants associated with NetSHy scores. This work demonstrates both the utility of a combined proteomic-genetic-network approach to identify novel proteins and their interactions involved in COPD phenotypes.

## Availability of Data and Materials

The SomaScan data supporting the conclusions of this article are available from the data coordinating centers of COPDGene and SPIROMICS respectively. The genomic data is available through TOPMed. The open-source code and reproducible analysis scripts can be accessed at https://github.com/KechrisLab/ProteinNetworks/.

## Consent for publication

Not applicable

## Authors’ Contributions

IK, TV, WL, EL, KP, KK analyzed and interpreted proteomics data. IK, EL, LV, NG performed QTL analyses. IK, TV, WL, KP, LV, BG, KK contributed to the writing of the initial draft. All authors participated in the reviewing and editing process. All authors have read and approved the final manuscript.

## Data Availability

The SomaScan data supporting the conclusions of this article are available from the data coordinating centers of COPDGene and SPIROMICS respectively. The genomic data is available through TOPMed.

https://github.com/KechrisLab/ProteinNetworks/

## Acknowledgements

Molecular data for the Trans-Omics in Precision Medicine (TOPMed) program was supported by the National Heart, Lung and Blood Institute (NHLBI). Genome Sequence data for "NHLBI TOPMed: COPDGene" (phs000951) was performed at Broad Genomics and the Northwest Genome Center at the University of Washington (NWGC) (HHSN268201500014C, 3R01HL089856-08S1). Core support including centralized genomic read mapping and genotype calling, along with variant quality metrics and filtering were provided by the TOPMed Informatics Research Center (3R01HL-117626-02S1; contract HHSN268201800002I). Core support including phenotype harmonization, data management, sample-identity QC, and general program coordination were provided by the TOPMed Data Coordinating Center (R01HL-120393; U01HL-120393; contract HHSN268201800001I). We gratefully acknowledge the studies and participants who provided biological samples and data for TOPMed.

The COPDGene project described was supported by Award Number U01 HL089897 and Award Number U01 HL089856 from the National Heart, Lung, and Blood Institute. The content is solely the responsibility of the authors and does not necessarily represent the official views of the National Heart, Lung, and Blood Institute or the National Institutes of Health. The COPDGene project is also supported by the COPD Foundation through contributions made to an Industry Advisory Board comprised of AstraZeneca, Boehringer Ingelheim, GlaxoSmithKline, Novartis, Pfizer, Siemens and Sunovion. A full listing of COPDGene investigators can be found at: http://www.copdgene.org/directory.

The authors thank the SPIROMICS participants and participating physicians, investigators, study coordinators, and staff for making this research possible. More information about the study and how to access SPIROMICS data is available at www.spiromics.org. The authors would like to acknowledge the University of North Carolina at Chapel Hill BioSpecimen Processing Facility (http://bsp.web.unc.edu/) and Alexis Lab (https://www.med.unc.edu/cemalb/facultyresearch/alexislab/) for sample processing, storage, and sample disbursements.

We would like to acknowledge the following current and former investigators of the SPIROMICS sites and reading centers: Neil E Alexis, MD; Wayne H Anderson, PhD; Mehrdad Arjomandi, MD; Igor Barjaktarevic, MD, PhD; R Graham Barr, MD, DrPH; Patricia Basta, PhD; Lori A Bateman, MS; Christina Bellinger, MD; Surya P Bhatt, MD; Eugene R Bleecker, MD; Richard C Boucher, MD; Russell P Bowler, MD, PhD; Russell G Buhr, MD, PhD; Stephanie A Christenson, MD; Alejandro P Comellas, MD; Christopher B Cooper, MD, PhD; David J Couper, PhD; Gerard J Criner, MD; Ronald G Crystal, MD; Jeffrey L Curtis, MD; Claire M Doerschuk, MD; Mark T Dransfield, MD; M Bradley Drummond, MD; Christine M Freeman, PhD; Craig Galban, PhD; Katherine Gershner, DO; MeiLan K Han, MD, MS; Nadia N Hansel, MD, MPH; Annette T Hastie, PhD; Eric A Hoffman, PhD; Yvonne J Huang, MD; Robert J Kaner, MD; Richard E Kanner, MD; Mehmet Kesimer, PhD; Eric C Kleerup, MD; Jerry A Krishnan, MD, PhD; Wassim W Labaki, MD; Lisa M LaVange, PhD; Stephen C Lazarus, MD; Fernando J Martinez, MD, MS; Merry-Lynn McDonald, PhD; Deborah A Meyers, PhD; Wendy C Moore, MD; John D Newell Jr, MD; Elizabeth C Oelsner, MD, MPH; Jill Ohar, MD; Wanda K O’Neal, PhD; Victor E Ortega, MD, PhD; Robert Paine, III, MD; Laura Paulin, MD, MHS; Stephen P Peters, MD, PhD; Cheryl Pirozzi, MD; Nirupama Putcha, MD, MHS; Sanjeev Raman, MBBS, MD; Stephen I Rennard, MD; Donald P Tashkin, MD; J Michael Wells, MD; Robert A Wise, MD; and Prescott G Woodruff, MD, MPH. The project officers from the Lung Division of the National Heart, Lung, and Blood Institute were Lisa Postow, PhD, and Lisa Viviano, BSN; SPIROMICS was supported by contracts from the NIH/NHLBI (HHSN268200900013C, HHSN268200900014C, HHSN268200900015C, HHSN268200900016C, HHSN268200900017C, HHSN268200900018C, HHSN268200900019C, HHSN268200900020C), grants from the NIH/NHLBI (U01 HL137880, U24 HL141762, R01 HL182622, and R01 HL144718), and supplemented by contributions made through the Foundation for the NIH and the COPD Foundation from Amgen; AstraZeneca/MedImmune; Bayer; Bellerophon Therapeutics; Boehringer-Ingelheim Pharmaceuticals, Inc.; Chiesi Farmaceutici S.p.A.; Forest Research Institute, Inc.; Genentech; GlaxoSmithKline; Grifols Therapeutics, Inc.; Ikaria, Inc.; MGC Diagnostics; Novartis Pharmaceuticals Corporation; Nycomed GmbH; Polarean; ProterixBio; Regeneron Pharmaceuticals, Inc.; Sanofi; Sunovion; Takeda Pharmaceutical Company; and Theravance Biopharma and Mylan/Viatris.

