## Supplementary Material for "Proteomic Networks and Related Genetic Variants Associated with Smoking and Chronic Obstructive Pulmonary Disease"

**Supplementary Methods**

Details on Matched Non-Hispanic White and African American Population

To create the matched dataset, all of the eligible AA participants were used to match the NHW group. For matching, the AA age was divided into quartiles: 44-54.7; 54.8-59.5; 58.5-63.9; and >63.9. The characteristics matched on were: sex, current smoking status, age category and COPD GOLD Stage. Because the AA participants were much younger and had greater than two times the number of current smokers; the matched dataset was modified in the following ways to refine matching. To improve matching on age and current smoking status, the younger two age categories and current smokers were deleted from the matched dataset and the total younger 2 age categories and all current smoker in the NHWs were added back to create the final dataset. This resulted in an improved match between the two groups for sex and current smoking status, but resulted in a difference in the number of observations included in the two populations.

Details on Proteomics Profiling

COPDGene was quantified with v4.0 and SPIROMICS v4.1 platforms. Both platforms had the same quality control applied. The SomaScan 4.0 platform uses 4,979 different SOMAmers (aptamers) to quantify 4,776 unique proteins with 4,720 unique Uniprot numbers. The SomaScan v4.1 platform includes all the SOMAmers as v4.0 with an additional 2,309 SOMAmers (total = 7,288). The common SOMAmers/proteins were used for replication. Results in the replication analysis were similar with or without the scale factor (also referred to as Version Lifting) to harmonize between the v4.0 and v4.1 platforms. We report results without the scale factor. At the COPDGene Visit 2, 6018 subjects were profiled using the SomaScan v4.0 platform, 815 were removed for the following reasons: 3 samples quality (small volume) or eligibility, 228 failed QC, 19 lung transplants, 98 potential mismatch of plasma sample labels [1], 402 never smoked cohort, and 65 missing data on matching variables. Following removal of these samples, 5203 subjects were available for matching on sex, smoking status, age, and COPD Gold stage (**Supplement Figure 1**). The final matched populations were 1660 non-Hispanic Whites and 1513 non-Hispanic Blacks (**Supplement Tables 1 and 2**). We used a coefficient of variation (CoV) threshold to filter out proteins with low variability (CoV < 0.7). After filtering, 2894 proteins remain for network inference.

The SPIROMICS Visit 1 had 2205 subjects with SomaScan 4.1 results were used to create a replication dataset. Four hundred thirteen observations were removed for the following reasons: 101 failed QC, 3 self-reported at Visit 1 having a history of lung volume reduction surgery, 3 with a potential mismatch of plasma sample labels,146 never smoked stratum, 160 self-identifying as being another race other than NHW or AA or self-reported Hispanic ethnicity (**Supplement Figure 2)**. The final replication population was 1792 subjects (1459 NHW and 333 AA) (**Supplement Table 1)**.

Univariate Analysis

We performed univariate correlation analysis to check the correlation between each of the 2894 proteins with respect to each phenotype using a Pearson’s correlation test. Both nominal and FDR-adjusted p-values are reported for each subnetwork (**Supplement Table 3**). Then we report the number of proteins in the subnetworks that are significant based on the FDR-adjusted p-value (**Table 2**).

Details on Network Analysis

*Network construction:* For all networks constructed, we obtained an adjacency matrix after extracting the canonical weights or PLS weights from SmCCA (continuous phenotype) or SPLS-DA (binary exposure) respectively, which represents the strength of connections between protein features. For SmCCA, the adjacency matrix is obtained by taking the outer product of the canonical weights. For SPLS-DA, often more than one component is used to achieve better prediction accuracy, and in this case, the feature importance score is obtained by taking the weighted sum of 3 component scores weighted by logistic regression coefficients after fitting the classification model in the second stage. Then the feature importance score is used to construct the adjacency matrix by taking the outer product between this score vector and itself. After the adjacency matrix is obtained using either of the two approaches, hierarchical clustering is implemented to partition proteins into different sub-network modules using dynamic tree cut [2].

*Sparsity penalties:* 5-fold cross-validation was performed to select sparsity penalty parameters for continuous traits. We perform 100 sub-sampling steps to stabilize the canonical weight by randomly selecting 70% of the features in the data. We performed a grid search of sparse penalty terms ranging from 0.1 (stringent) to 0.5 (relaxed) with a step size of 0.1. We used cross-validation in this case to validate the prediction error corresponding to each penalty term. The criterion used to select the best penalty term was defined as

$\min\frac{\left| test cc - train cc \right|}{\left| test cc \right|}$,

where cc is the canonical correlation, which aimed to (1) minimize the canonical correlation discrepancy between the training set and the testing set, and (2) maximize the testing canonical correlation. For the binary scenario (smoking), it was the reverse of SmCCA, using PLS-DA such that the sparse penalty terms ranged from 0.1 (relaxed) to 0.9 (stringent) with a step size of 0.1. To evaluate the best penalty term through cross-validation, we maximized the test accuracy of predicting smoking status.

*Network trimming:* The following procedure was used for trimming: (1) For each network module, we run the PageRank algorithm [3] on the complete network and obtain the PageRank score for each protein, then (2) we choose a subset of the 10 top proteins by the PageRank score, and treat them as the baseline network (3) we increase the network size by one (i.e., include one new protein into the network based on PageRank score), and calculate the NetSHy summarization score and (4) calculate two evaluation metrics to decide whether this new network is accepted. This process continues until the final protein from the original sub-network is considered. The two evaluation metrics are (a) correlation between summarization score and original phenotype (within 90% range of the maximum correlation between summarization score and phenotype), and (b) correlation between the summarization of the current network size and that of baseline network size (>0.80). If these criteria are not met, then the trimmed network is returned.

*Network visualization with subnetwork edge filtering:* We use Cytoscape (version 3.9.0) [4] to visualize the proteomics network modules. To simplify the steps of importing the adjacency matrices to Cytoscape, we utilize the R package RCy3 [5] to directly communicate between R studio and Cytoscape. The final subnetwork obtained from the node trimming is densely connected, which may contain false positive edges. Therefore, for visualization purposes, we calculate the correlation between each pair of nodes, and after trying different correlation thresholds and checking the network, we use 0.05 as the correlation threshold to filter out edges that connect nodes that are less correlated.

*Statistical test for comparing subnetworks:*

We represent each identified phenotype-specific subnetwork with adjacency matrices $A^{NHW}$ and $A^{AA}$ of size $M\times M$, for the two populations NHW and AA, respectively. We denote $V^{NHW}$ and $V^{AA}$ as vectors of the lower triangles of the corresponding $A^{NHW}$ and $A^{AA}$. The difference between $V^{NHW}$ and $V^{AA}$, denoted by $D$, can be computed as follows:

$$D=PND\left( {V^{NHW},V}^{AA}, p=6 \right)=\left( \frac{1}{K}\sum_{k=1}^{K} \left| V_{k}^{NHW}-V_{k}^{AA} \right|^{6} \right)^{1/6}$$

with $K=\frac{M(M-1)}{2}$.

The closer the value of $D$ to 0, the more similar the two subnetworks. Given that $D\geq0$, we are interested in testing the following hypotheses: $H_{0}: D=0$ vs. $H_{A}:D>0$. Following Arbet et al. [6], we use a non-parametric permutation method to derive the sampling distribution under the null hypothesis. The following procedure is conducted to calculate the permutation p-values:

For $b=1, 2, \ldots, B$ ($B$ is total number of permutations):

i. Using the observed vectors $V^{NHW}$ and $V^{AA}$, we disrupt their pair order relationship by fixing $V^{NHW}$ and randomly permuting $V^{AA}$ to create $V^{AA}(b)$.
ii. Calculating the permuted test statistic $D(b)$ based on $V^{NHW}$ and $V^{AA}(b)$.
iii. Computing the permutation p-value $=\frac{\sum_{b=1}^{B} \left[ I\left( \left| D\left( b \right) \right|>\left| D \right| \right) \right]+1}{B+1}$, with $I(.)$ as an indicator function.

P-values that are smaller than the significance level α correspond to rejecting the null hypothesis at α level, indicating that the two comparing subnetworks are different.

*Network projection:*

We also performed complementary network comparison called network projection which takes a subnetwork (adjacency matrix) in one group and protein abundance in another. This is useful in settings when network construction was only performed in one cohort (COPDGene) but not another (SPIROMICS). We also use it to compare the two groups in COPDGene to check for consistency with the statistical test of adjacency matrices.

*Cross-race-group projection:* We use a subnetwork derived from NHW population, denoted by $A^{NHW}$, to generate a corresponding Laplacian matrix $L^{NHW}$. We then impose $L^{NHW},$ which captures the network connectivity from the NHW population, onto the proteomics profile of the AA population, denoted by $X^{AA}$, in order to compute the NetSHy summarization scores as in [7]. We refer to these as NHW projection scores which are then used to calculate the correlation with the phenotype of interest. Similarly, we project the AA network (captured in $L^{AA}$) onto the NHW population ($X^{NHW}$) to obtain the AA projection scores. This process is repeated for each phenotype or exposure. The projection scores are correlated with the corresponding phenotype/exposure, denoted as projection correlations which are then used to statistically compare with the NetSHy correlations. A bootstrap sampling procedure described below was used to construct 95% confidence intervals.

*Bootstrap sampling:* While projecting subnetwork $L^{NHW}$onto the proteomics data $X^{AA}$, we implement bootstrap sampling to generate a distribution for the projection correlations. In particular, we generate 1000 bootstrap samples with replacement from $X^{AA}$, denoted by $X_{\left( u \right)}^{AA}$. By keeping $L^{NHW}$ fixed, we compute the NetSHy summarization scores and the corresponding projection correlations (denoted by $\rho_{\left( u \right)}$) for each $u^{th}$ bootstrap sample. We obtain the 95% confidence interval for the projection correlation, which ranges from the ${2.5}^{th}$ and ${97.5}^{th}$ percentiles of the bootstrap correlations $\rho_{\left( u \right)}$.

*Cross-cohort projection:* We further extend our projection procedure in SPIROMICS, which is another independent TOPMed cohort. Note that we will not have networks in SPIROMICS to use the PND6 statistical testing approach described above, but can still project onto the available SPIROMICS proteomic data. Specifically, we keep the NHW population as a fixed reference and project each phenotype-specific subnetwork derived from COPDGene onto the proteomics data in SPIROMICS to obtain cross-cohort NHW projection scores. Similar procedure is performed while keeping the AA population as a reference to extract cross-cohort AA projection scores. The resulting cross-cohort projection scores are correlated with their respective phenotypes/exposure and exposure to generate cross-cohort correlations. These correlations are compared with the original NetSHy correlations to assess the replicability of the subnetworks across independent cohorts.

COPDGene Whole-Genome Sequencing (WGS)

Briefly, samples were sequenced with 150 bp paired-end reads for a target depth of ≥ 30 with harmonized protocols at two NHLBI-sequencing centers (the Broad Institute of MIT and Harvard and the University of Washington Northwest Genomics Center), using Illumina HiSeq X platform (Illumina, San Diego, California). A common pipeline was used across centers to align reads to GRCh38 and perform genotype calling. We obtained Freeze 10 (Jan 2022) WGS data. The generation of Freeze 9 WGS data has been previously described [8] and Freeze 10 is similar with additional samples. Variants from WGS data were filtered by a minor allele frequency (MAF) ≥ 5%, and a standard Bonferroni genome-wide significance threshold of 5x10^-8^ was used for each phenotype.

Colocalization in Single Protein Quantitative Trait Loci Analysis Comparison

We also tested the hypothesis that the network quantitative trait loci (nQTLs) were driven by single protein quantitative trait loci (pQTLs) in a two-step analysis. First, we used a Bayesian implementation to formally test for colocalization between nQTLs and single-protein genetic associations for each NetSHy score, each trait, and each population in which these signals co-occurred on the same chromosome, using R package *coloc* v. 5.1.0.1 [9]. For instances of probable colocalization (defined as a posterior probability for the shared variant hypothesis (H4) ≥ 0.9), we performed a conditional nQTL analysis in which we further adjust for the single-protein abundance values of the protein with a colocalizing genetic signal to quantify whether the nQTL association is independent of the single-protein genetic association.

**Supplement Tables**

| **Supplement Table 1. Characteristics of NHW matched and not matched** | | | |
| --- | --- | --- | --- |
| **Characteristics** | **NHW**  **N=1660** | **NHW Not Matched**  **N=2030** | **p-value** |
| ***Demographics*** |  |  |  |
| Age (yr) mean (SD) | 62.9 (8.0) | 71.5 (6.3) | <0.001 |
| Males n(%) | 790 (47.6%) | 1089 (53.7%) | <0.001 |
| Females n(%) | 870 (52.4%) | 941 (46.3%) |  |
| ***Smoking Exposure*** |  |  |  |
| Smoking Status n(%): Former | 647 (39.0%) | 2030 (100%) | <0.001 |
| Current | 1013 (61.0%) | 0 (0.0%) |  |
| Pack-years median(IQR) | 42.3 (25.5) | 42.0 (31.5) | 1.0 |
| ***Clinical*** |  |  |  |
| BMI kg/m^2^ (mean(SD)) | 28.4 (6.3) | 29.2 (5.8) | <0.001 |
| COPD GOLD Stages n(%): PRISm | 209 (12.6%) | 180 (8.9%) | <0.001 |
| GOLD 0 Smoker Controls | 678 (40.8%) | 817 (40.2%) |  |
| GOLD 1 | 158 (9.5%) | 240 (11.8%) |  |
| GOLD 2 | 370 (22.3%) | 427 (21.0%) |  |
| GOLD 3 | 185 (11.1%) | 247 (12.2%) |  |
| GOLD 4 | 60 (3.6%) | 119 (5.9%) |  |
| ***Pulmonary Function*** (mean(SD)) |  |  |  |
| FEV1 Post (liter/sec) | 2.3 (0.9) | 2.0 (0.8) | <0.001 |
| FEV1 Percent Predicted | 76.8 (23.4) | 76.7 (26.2) | 0.91 |
| FEV1/FVC | 0.67 (0.15) | 0.60 (0.20) | <0.001 |
| FVC (liter) | 3.3 (1.0) | 3.1 (0.9) | <0.001 |
| ***Emphysema*** |  |  |  |
| Emphysema (% LAA < -950 HU) median(IQR) | 1.5 (4.4) | 2.6 (8.1) | <0.001 |
| PD15_adj_ (g/L) | 87.0 (24.0) | 77.9 (22.7) | <0.001 |
| ***Blood Cell Count*** (mean (SD)) |  |  |  |
| Platelets (K/μL) | 242.3 (70.4) | 233.7 (64.0) | 0.001 |
| WBC (K/μL) | 7.6 (2.1) | 7.2 (2.4) | <0.001 |
| ^+^P-values comparison of the two populations using a chi square test for categorical data (Fisher exact if cell count <5), 2-sample t-tests for normally distributed continuous variables and Wilcoxon Rank Sums tests for non-normal continuous variables  NHW: non-Hispanic White; AA; PRISm: Preserved ratio impaired spirometry; FEV_1_: Forced expiratory volume in 1 second; FEV_1_ percent predicted: Percentage of lung capacity expelled in 1 second; FEV_1_/FVC: Ratio of the forced expiratory volume in one second to the forced vital capacity of the lung. | | | |

| **Supplement Table 2. Characteristics of SPIROMICS and COPDGene Study populations** | | | |
| --- | --- | --- | --- |
|  | SPIROMICS  N=1792 | COPDGene  N=3173 | p-value* |
| ***Demographics*** |  |  |  |
| Age (yr) mean (SD) | 64.1 (8.7) | 61.6 (7.7) | <0.001 |
| Non-Hispanic White n(%) | 1459 (81.4) | 1660 (52.3) | <0.001 |
| African American n(%) | 333 (18.6) | 1513 (47.7) |  |
| Males n(%) | 967 (54.0) | 1535 (48.4) | <0.001 |
| Females n(%) | 825 (46.0) | 1638 (51.6) |  |
| ***Smoking Exposure*** |  |  |  |
| Smoking Status n(%): Former | 684 (38.8) | 1995 (62.9) | <0.001 |
| Current | 1081 (61.2) | 1178 (37.1) |  |
| Pack-years median(IQR) | 45.00 (27.0) | 39.00 (25.3) | <0.001 |
| ***Clinical*** |  |  |  |
| BMI (mean (SD)) | 27.8 (5.3) | 28.9 (6.7) | <0.001 |
| COPD GOLD Stages n(%): PRISm | 46 (2.6) | 465 (14.7) | <0.001 |
| GOLD 0 Smoker Controls | 534 (29.9) | 1405 (44.3) |  |
| GOLD 1 | 269 (15.0) | 263 (8.3) |  |
| GOLD 2 | 535 (29.9) | 617 (19.4) |  |
| GOLD 3 | 282 (15.8) | 314 (9.9) |  |
| GOLD 4 | 122 (6.8) | 109 (3.4) |  |
| ***Pulmonary Function*** (mean(SD)) |  |  |  |
| FEV_1_ (liter) | 2.1 (0.9) | 2.2 (0.8) | <0.001 |
| FEV_1_ Percent Predicted | 72.7 (26.3) | 78.3 (23.6) | <0.001 |
| FEV_1_/FVC | 0.60 (0.17) | 0.69 (0.14) | <0.001 |
| FVC (liter) | 3.4 (1.02) | 3.1 (0.97) | <0.001 |
| ***Emphysema*** |  |  |  |
| Emphysema (% LAA < -950 HU) Median(IQR) | 3.53 (10.2) | 1.20 (4.1) | <0.001 |
| PD15_adj_ (g/L) (mean(SD)) | 80.8 (26.9) | 90.7 (26.3) | <0.001 |
| ***Blood Cell Counts*** (mean (SD)) |  |  |  |
| Platelets (K/μL) | 247.2 (68.6) | 242.1 (71.6) | 0.014 |
| WBC (K/μL) | 7.11 (2.2) | 7.15 (2.3) | 0.536 |
| ^+^P-values comparison of the two populations using a chi square test for categorical data, 2-sample t-tests for normally distributed continuous variables and Wilcoxon Rank Sums tests for non-normal continuous variables  NHW: non-Hispanic White; AA: African American; PRISm: Preserved ratio impaired spirometry; FEV_1_: Forced expiratory volume in 1 second; FEV_1_/FVC: Ratio of the forced expiratory volume in one second to the forced vital capacity. | | | |

S**upplemental Table 3:** See attached spreadsheet.

**Supplemental Table 4a: Summary results of network projections using subnetworks associated with FEV_1_ across NHW and AA, in two independent cohort studies COPDGene (C) and SPIROMICS (S).**

| NHW Data | | | | | |
| --- | --- | --- | --- | --- | --- |
|  | Network | Data | Component | | |
|  |  |  | 1 | 2 | 3 |
| Original | C-NHW | C-NHW | 0.137 | 0.154 | 0.073 |
| Across populations | C-AA | C-NHW | 0.060  (0.015, 0.108) | 0.032  (0.002, 0.083) | **0.045**  **(0.003, 0.092)** |
| Across cohorts | C-NHW | S-NHW | **0.080**  **(0.022, 0.143)** | **0.165**  **(0.110, 0.215)** | **0.052**  **(0.002, 0.116)** |
| Across populations & cohorts | C-AA | S-NHW | **0.087**  **(0.038, 0.139)** | **0.105**  **(0.047, 0.158)** | **0.082**  **(0.013, 0.139)** |
| AA Data | | | | | |
|  | Network | Data | Component | | |
|  |  |  | 1 | 2 | 3 |
| Original | C-AA | C-AA | 0.144 | 0.032 | 0.028 |
| Across populations | C-NHW | C-AA | 0.022  (0.001, 0.081) | **0.087**  **(0.029, 0.155)** | **0.026**  **(0.002, 0.111)** |
| Across cohorts | C-AA | S-AA | **0.036**  **(0.002, 0.160)** | **0.145**  **(0.021, 0.280)** | 0.132  (0.015, 0.277) |
| Across populations & cohorts | C-NHW | S-AA | **0.069**  **(0.004, 0.180)** | **0.003**  **(0.002, 0.218)** | **0.144**  **(0.006, 0.240)** |

Original correlations represent correlations between NetSHy scores and FEV_1_ in a population within the same cohort. The remaining correlations are calculated through network projections across populations and/or cohorts. The 95% bootstrap confidence intervals are recorded in parentheses. Values in bold are when the original correlations (first row in each sub table) fall within the confidence interval, indicating replication.

**Supplemental Table 4b: Summary results of network projections using subnetworks associated with %LAA950 across two populations NHW and AA, in two independent cohort studies COPDGene (C) and SPIROMICS (S).**

| NHW Data | | | | | |
| --- | --- | --- | --- | --- | --- |
|  | Network | Data | Component | | |
|  |  |  | 1 | 2 | 3 |
| Original | C-NHW | C-NHW | 0.144 | 0.091 | 0.014 |
| Across populations | C-AA | C-NHW | **0.101**  **(0.059, 0.146)** | **0.004**  **(0.001, 0.092)** | **0.073**  **(0.01, 0.129)** |
| Across cohorts | C-NHW | S-NHW | **0.113**  **(0.059, 0.163)** | **0.005**  **(0.001, 0.065)** | **0.017**  **(0.001,0.085)** |
| Across populations & cohorts | C-AA | S-NHW | **0.063**  **(0.01, 0.119)** | **0.126**  **(0.075, 0.184)** | **0.095**  **(0.01, 0.135)** |
| AA Data | | | | | |
|  | Network | Data | Component | | |
|  |  |  | 1 | 2 | 3 |
| Original | C-AA | C-AA | 0.077 | 0.067 | 0.118 |
| Across populations | C-NHW | C-AA | **0.095**  **(0.044, 0.144)** | **0.060**  **(0.004,0.123)** | 0.043  (0.004,0.098) |
| Across cohorts | C-AA | S-AA | **0.069**  **(0.003, 0.173)** | **0.166**  **(0.031, 0.258)** | **0.007**  **(0.004,0.211)** |
| Across populations & cohorts | C-NHW | S-AA | **0.071**  **(0.005, 0.174)** | **0.120**  **(0.006, 0.239)** | **0.035**  **(0.002,0.199)** |

Original correlations represent correlations between NetSHy scores and %LAA950 in a population within the same cohort. The remaining correlations are calculated through network projections across populations and/or cohorts. The 95% bootstrap confidence intervals are recorded in parentheses. Values in bold are when the original correlations (first row in each sub table) fall within the confidence interval, indicating replication.

| **Supplement Table 5. Colocalization of NetSHy and single protein GWAS results.** | | | | | | | | | | |
| --- | --- | --- | --- | --- | --- | --- | --- | --- | --- | --- |
| **Population** | **Phenotype** | **Trait 1** | **Trait 2** | **Region** | **# SNPs** | **PP (H0)** | **PP (H1)** | **PP (H2)** | **PP (H3)** | **PP (H4)** |
| NHW | %LAA950 | NetSHy3 | Cadherin 17 | chr9:132423455-134176000 | 4,766 | 1.22E-29 | 3.09E-25 | 2.40E-07 | 5.09E-03 | 0.995 |
| NHW | Smoking | NetSHy1 | Alkaline phosphatase placental type | chr2:230978674-232685293 | 3,153 | 1.71E-32 | 1.83E-16 | 3.17E-18 | 0.0329 | 0.967 |
| NHW | Smoking | NetSHy1 | Alkaline phosphatase placental like 1 | chr2:230978674-232685293 | 3,153 | 2.66E-29 | 2.85E-13 | 2.85E-18 | 0.0296 | 0.97 |
| AA | Smoking | NetSHy1 | Alkaline phosphatase placental type | chr2:231720130-232500432 | 1,567 | 1.09E-09 | 3.56E-07 | 4.83E-06 | 5.76E-04 | 0.999 |
| NHW | FEV_1_ | NetSHy2 | Left right determination factor 2 | chr1:224750818-226623159 | 3,662 | 2.46E-99 | 4.63E-84 | 6.40E-17 | 0.12 | 0.88 |
| NHW | %LAA950 | NetSHy3 | Cadherin 17 | chr19:46646825-52482037 | 13,300 | 1.16E-26 | 1.31E-23 | 8.87E-04 | 0.999 | 4.25E-05 |
| NHW | Smoking | NetSHy1 | Gastrokine 2 | chr2:230978674-232685293 | 3,153 | 7.87E-17 | 0.843 | 1.20E-17 | 0.128 | 0.0293 |

The table shows colocalization tests of overlapping genetic associations between protein sub-networks (Trait 1) and single proteins in the corresponding sub-network (Trait 2). The coordinates to each region are listed as the chromosome, followed by the start and end positions (reference GRCh38). # SNPs refers to the number of polymorphic sites identified in each region. The posterior probabilities of the colocalization test are listed (P0-P4), where H0 is the hypothesis that none of the variants are causal, H1: causal variant for trait 1 only, H2: causal variant for trait 2 only, H3: two distinct causal variants, and H4: one common causal variant (colocalized signals). A posterior probability (PP) greater than 0.9 indicates probable hypothesis. AA: African American. NHW: Non-Hispanic white.

**Supplement Figures**

**Year 5 (Phase 2)**

**EDTA Plasma**

**N=6018**

**N=5787**

228 Failed normalization acceptance criteria

3 Participant due to sample quality or eligibility

19 Lung transplants

98 potential plasma tube ID label error

**Year 5 (Phase 2)**

**Final Dataset**

**N=5670**

402 Never smoked cohort

65 Data missing on matching variables

**Available for Matching**

**N=5203**

2030 non-Hispanic White former smoker not matched

**Matched**

Sex, CurrentSmoking Status, Age, COPD GOLD Stage

**N=3173**

**non-Hispanic White**

**N=1660**

**non-Hispanic AA**

**N=1513**

**Supplement Figure 1.** Consort diagram of the COPDGene non-Hispanic White and African American (AA) cohorts. SomaLogic analyzed 6018 EDTA plasma samples. After removing results from samples with poor quality, failed normalization acceptance criteria, and participants that were found to not be eligible to be in the study, left 5787 eligible samples. Of those, 19 were participants who had reported having a lung transplant or lung volume reduction surgery, and 98 that were found to have a potential plasma tube ID labeling error. There were 5670 participants with SomaScan and clinical data. The study focused on those who had ever smoked; therefore, the never smoked cohort was removed as well as the 65 subjects that had missing values on any of the matching variables (sex, current smoking status, age, and COPD GOLD Stage). Of the 3690 non-Hispanic White population eligible for matching, 2030 that were former smokes that were not matched, their characteristics are in the Supplement Table 1.

**Visit 1**

**EDTA Plasma**

**N=2205**

**N=2101**

101 Failed normalization acceptance criteria

3 LVRS

3 potential plasma tube ID label error

**Visit 1**

**Final Dataset**

**N=2098**

146 Never smoked stratum

**Final SPIROMICS SomaScan Dataset**

**N=1792**

**non-Hispanic White**

**N= 1459**

**non-Hispanic AA**

**N=333**

160 Races other than NHW or AA or Hispanic ethnicity

**Supplement Figure 2**. Consort diagram of the SPIROMICS dataset creation. SomaLogic analyzed 2205 EDTA plasma samples. After removing results from samples that failed normalization acceptance criteria, left 2101 eligible samples. Of those, 3 were participants who had reported having a history of Lung Volume Reduction Surgery (LVRS) at Visit 1, and 3 that were found to have a potential plasma tube ID labeling error. There were 2098 participants with SomaScan and clinical data. To create an ever-smoked COPD cohort similar to COPDGene the never-smoked stratum was removed, as well as the participants how reported another race other than NHW or AA or being Hispanic. Characteristics of the SPIROMICS replication population are in Supplement Table 2.

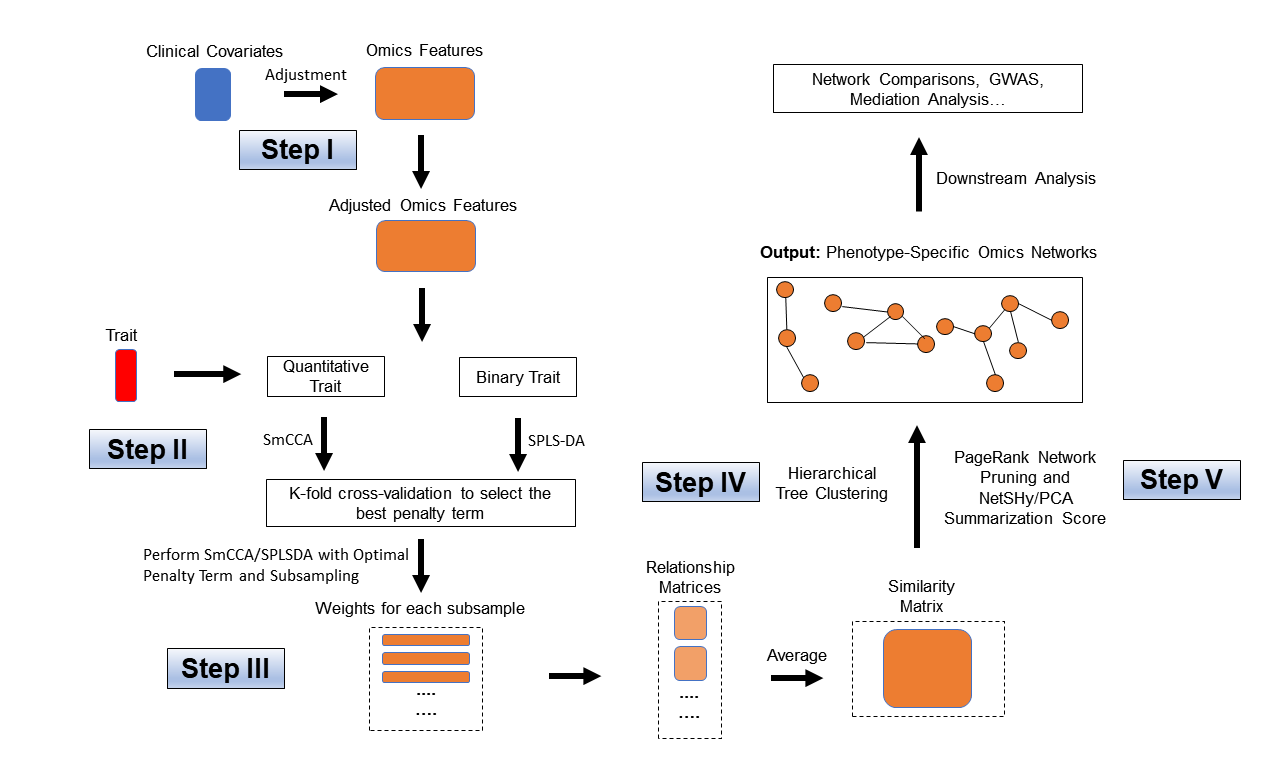

**Supplement Figure 3: Workflow of Single-Omics SmCCNet.** The workflow of Single-omics SmCCNet that consists of 5 steps. Step I: Effect of selected clinical covariates is regressed out from each molecular feature; Step II: Perform Cross-validation to select the best penalty term for Sparse multiple Canonical Correlation Analysis (SmCCA) for quantitative phenotype or Sparse Partial Least Squared Discriminant Analysis (SPLS-DA) for binary phenotype; Step III: Run SmCCA or SPLS-DA with selected penalty term with subsampling the extract robust canonical weights, and construct similarity matrix for molecular features; Step IV: Perform hierarchical clustering to partition molecular features into different subnetwork modules; Step V: Perform network pruning with NetSHy summarization score and PageRank algorithm to prune each subnetwork.

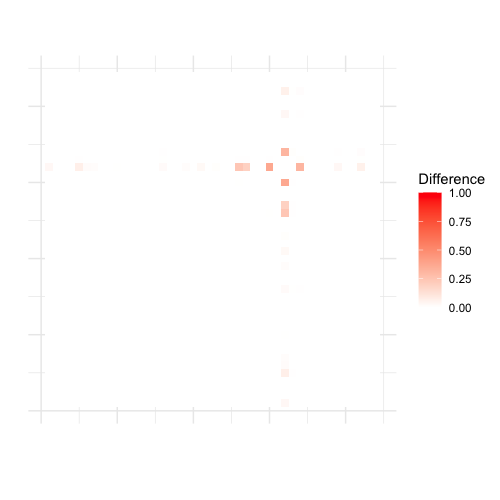

**Supplement Figure 4a: Heatmaps for differences between networks associated with smoking exposure between NHW and AA populations.** Using a network derived from NHW as an anchor (**left**), we calculate corresponding edge-wise differences between the two populations to construct a heatmap. Similarly, using a network from AA population as an anchor (**right**), we obtain the heatmap for the corresponding edge-wise differences between the two populations. The deeper red indicates a bigger difference.

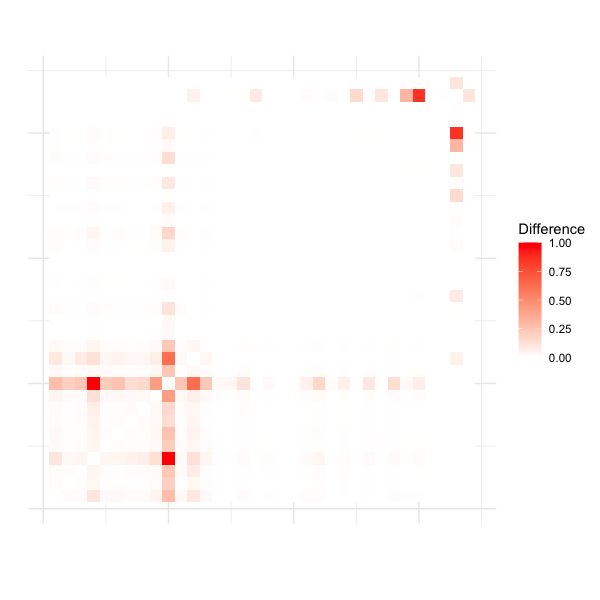

**Supplement Figure 4b: Heatmaps for differences between networks associated with FEV1 between NHW and AA populations.** Using a network derived from NHW as an anchor (**left**), we calculate corresponding edge-wise differences between the two populations to construct a heatmap. Similarly, using a network from AA population as an anchor (**right**), we obtain the heatmap for the corresponding edge-wise differences between the two populations. The deeper red indicates a bigger difference.

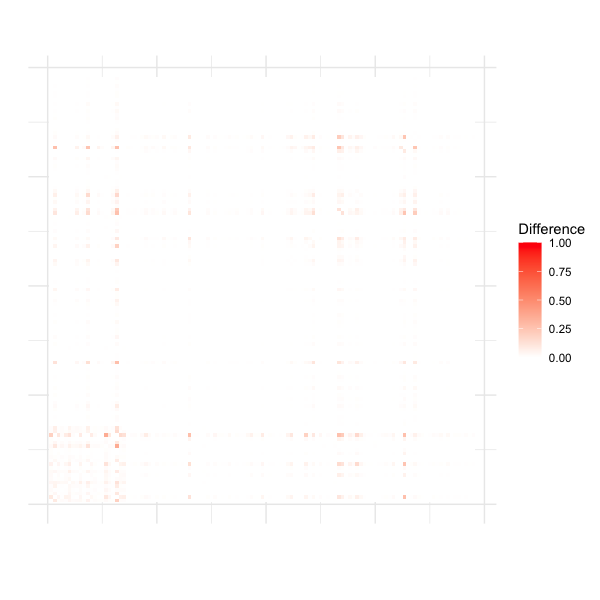

**Supplement Figure 4c: Heatmaps for differences between networks associated with EMP between NHW and AA populations.** Using a network derived from NHW as an anchor (**left**), we calculate corresponding edge-wise differences between the two populations to construct a heatmap. Similarly, using a network from AA population as an anchor (**right**), we obtain the heatmap for the corresponding edge-wise differences between the two populations. The deeper red indicates a bigger difference.
